# Integrated Metabolomics Identifies Mitochondrial Dysfunction, Oxidative Membrane Remodeling, and Reduced Lipophilic Antioxidant Defenses in First-Episode Simple Dysmood Disorder

**DOI:** 10.64898/2026.11.01.26366481

**Authors:** Visuthsiri Sirivatanapa, Pannipa Janta, Asara Vasupanrajit, Rossarin Tansawat, Andre F Carvalho, Abbas F Almulla, Chavit Tunvirachaisakul, Michael Maes

## Abstract

**Background:** Metabolomic abnormalities in major depression have largely been investigated at the level of individual metabolites. We hypothesized that coordinated metabolic pathways, rather than isolated biomarkers, characterize first-episode simple dysmood disorder (FE-SDMD).

**Methods:** Untargeted LC-MS metabolomics was performed in plasma from 44 FE-SDMD patients and 44 healthy controls. Differential metabolites were identified using Wilcoxon tests with false discovery rate correction and integrated with complementary machine-learning approaches. Curated metabolites were grouped into biologically informed functional modules, validated using principal component analysis, and examined in relation to clinical phenome dimensions, adverse childhood experiences (ACEs), and negative life events (NLEs).

**Results:** Of 450 detected metabolites, 298 remained significant after FDR correction and 240 additionally met fold-change criteria. Complementary feature-selection methods yielded a consensus panel of 20 biologically relevant metabolites that discriminated FE-SDMD from controls across multivariate and machine-learning analyses. Six functional modules revealed increased oxidative membrane remodeling, gut–liver dysfunction, mitochondrial bioenergetic dysfunction, reduced lipophilic antioxidant defenses and glucocorticoid metabolism, and altered dietary phytochemical metabolism. ACEs, particularly emotional neglect, and NLEs were associated with greater abnormalities across all modules. Gut–liver and mitochondrial dysfunction, oxidative stress, NLEs, and/or ACEs explained substantial variance in overall depression severity (66%), physiosomatic symptoms (41%), and suicidal ideation (43%).

**Conclusions:** Functional metabolic modules provide a biologically interpretable systems-biology framework linking metabolomic abnormalities, psychosocial adversity, and clinical phenotypes, supporting the neuroimmune–metabolic–oxidative (NIMETOX) model of depression.

## 1. Introduction

Major depressive disorder (MDD) is increasingly conceptualized as a disorder involving interconnected neuroimmune, metabolic, and oxidative stress (NIMETOX) pathways^1^. Within this framework, the central neuropathology and clinical phenome of MDD are considered to arise, at least partly, from systemic NIMETOX dysregulation and its interactions with brain function^1^. Over the past decades, extensive evidence has implicated immune activation, metabolic dysfunction, hypothalamic–pituitary–adrenal (HPA) axis dysregulation, oxidative and nitrosative stress (ONS), gut microbial dysbiosis, neurochemical alterations, impaired neural connectivity, and increased neurotoxicity in the pathophysiology and clinical expression of MDD^1–12^. However, despite extensive evidence for individual NIMETOX abnormalities, considerably less is known about how these alterations are organized into coordinated functional metabolic systems, how these systems interact with psychosocial adversity, and how they map onto specific dimensions of the depressive phenome.

Metabolomics provides an attractive approach for examining the crosstalk among NIMETOX pathways because it measures downstream biochemical products that are proximal to the clinical phenotype and integrate the influences of genetic background, environmental exposures, NIMETOX pathways, and the gut microbiome. Consequently, the metabolome provides a dynamic snapshot of ongoing biochemical activity and its interactions with environmental and host-related factors. Although previous metabolomic studies have identified numerous dysregulated metabolites in MDD^13–16^, findings have often been inconsistent across studies, insufficiently replicated, and difficult to integrate into coherent functional and systems biology pathways.

Previous metabolomic studies have largely interpreted MDD through individual metabolites identified using volcano plots, univariate tests, and variable importance in projection (VIP) scores. Although informative, such approaches may insufficiently capture the systems biology of the metabolome. Metabolites are highly interconnected and co-regulated, with metabolic pathways overlapping and sharing substrates and products. Consequently, single-metabolite approaches may yield fragmented and poorly reproducible biological interpretations while obscuring coordinated metabolic disturbances relevant to the clinical phenome.

A systems-biology alternative is to integrate functionally related and co-regulated metabolites into biologically informed metabolic modules^1,17,18^. Such modules may capture different functions including mitochondrial bioenergetics, oxidative membrane remodeling, gut– liver metabolism, and glucocorticoid regulation. This approach is particularly relevant to NIMETOX because immune-inflammatory, metabolic, redox, and neuroendocrine pathways are reciprocally interconnected and therefore predict coordinated rather than isolated abnormalities^1,19^.

Supporting this approach, deep metabolomic phenotyping of MDD identified six functional domains derived from 16 metabolites that explained 72.5% of the variance in overall severity and classified patients with 97.5% accuracy, whereas individual metabolites showed substantially weaker performance^1,20^. Moreover, lipotoxicity, including ceramide and diacylglycerol excess, and increased oxidative and nitrosative stress are detectable in first-episode (FE) patients without metabolic syndrome^18^, suggesting that these abnormalities are not restricted to chronic or metabolically compromised MDD.

Two important gaps remain. First, module-based metabolomics has focused mainly on chronic, recurrent, or hospitalized MDD, where illness burden and metabolic comorbidity may influence the metabolome^17,20^. Whether comparable abnormalities occur in FE simple dysmood disorder (SDMD) remains unknown; systems-level evidence is limited to our lipidomic study of this cohort^18^. Second, no study has integrated machine-learning feature selection, biological metabolite curation, functional module construction, and modelling of their relationships with dimensional phenotypes and psychosocial adversity.

Therefore, the present study aimed to (1) identify robust metabolomic features of FE-SDMD; (2) construct biologically informed functional modules from curated metabolites; (3) determine their associations with key depression domains including severity, physiosomatic symptoms, suicidal behaviors, insomnia, executive functions, and psychosocial adversity; and (4) develop an integrated systems-biology model of FE-SDMD.

## 2. Materials and Methods

This study used plasma samples and clinical data collected in a prior study with collection period between November 2021 and February 2023^21^. All samples and their associated clinical and biomarker data used in the present study were de-identified. The present study was approved by the Institutional Review Board (IRB) of the Faculty of Medicine, Chulalongkorn University (IRB No. 351/63 and IRB No. 0540/68, respectively).

### 2.1. Participants

The study comprised 88 Thai-speaking university students, including men and women aged 18–35 years and enrolled across different faculties of Chulalongkorn University, Bangkok, Thailand. The clinical group consisted of 44 students receiving outpatient care at the Department of Psychiatry, King Chulalongkorn Memorial Hospital, who met DSM-5 criteria for a first episode of major depressive disorder (MDD), as established by a senior psychiatrist, and fulfilled the research criteria for FE-SDMD. FE-SDMD is a research-defined phenotype nested within DSM-5 MDD that was developed using clinical, recurrence-of-illness (ROI), and biological data to identify an early and relatively uncomplicated form of MDD^22^; it is not an official DSM-5 or ICD diagnostic category. Patients were selected from 79 consecutively recruited MDD outpatients, of whom 35 were excluded because of a previous MDD episode, major dysmood disorder (MDMD), or metabolic syndrome (MetS). The healthy control (HC) group comprised 44 university students without MDD. Both groups were matched for age, sex, and body mass index (BMI). Exclusion criteria for study participation included MetS, pregnancy or breastfeeding, endocrine, autoimmune or other immune disorders, neurological or cardiovascular disorders, severe suicidal ideation, and other DSM-IV-TR axis 1 major psychiatric disorders. Additional exclusion criteria for controls were a lifetime history of MDD and dysthymia, and a family history of MDD, bipolar disorder, schizophrenia and suicide.

### 2.2. Data Collection and Clinical Assessments

A screening structured interview was completed by a trained clinical psychologist. The diagnosis of major psychiatric disorders (axis 1) was made using DSM-5 criteria (MDD, bipolar disorder, anxiety disorders, psychotic disorders, eating disorders, and substance used disorders) and The Mini International Neuropsychiatric Interview (M.I.N.I.)^23^. In addition, MDD was diagnosed using DSM-5 criteria using the Structured Interview according to DSM-5 (SCID).

All participants completed psychiatric assessments including severity of anxiety, depression, insomnia, suicidality, and stressful events (instruments and abbreviations in Supplementary Information, Methods). The first PC of total HAM-D, BDI-II, STAI, ISI, and current SB scores were used to capture overall severity of depression (OSOD)^21^.

MetS diagnosis was made according to the Joint Scientific Statement of the American Heart Association and the National Heart, Lung, and Blood Institute criteria^24^ (Supplementary Information, Methods). Potential effects of metabolic abnormalities below the diagnostic threshold for MetS were additionally controlled using two indices: the number of individual MetS components present as an ordinal measure of subclinical metabolic burden, and a dichotomous indicator distinguishing participants with no MetS components from those exhibiting at least one component.

### 2.3. Assays

All participants fasted for 8 hours prior to blood collection. Blood samples were obtained at 7:30 a.m. and stored at −80. Full information about data collection and the study protocol is available in our earlier publication^21^ (sample preparation, chromatography, mass spectrometry, and data processing in Supplementary Information, Methods).

Features annotated as unknown compounds were excluded from further analysis. All annotated metabolites were manually verified against HMDB records and published literature to confirm their classification as endogenous human plasma metabolites.

### 2.4. Statistical analysis

#### 2.4.1. Classical Statistical Analyses

Group differences in metabolomic variables were assessed using the Wilcoxon rank-sum test, whereas continuous demographic, clinical, and metabolomic variables were analyzed using analysis of variance (ANOVA) or covariance (ANCOVA), as appropriate. Correction for multiple comparisons was performed using the false discovery rate (FDR). The a priori sample size estimation is reported in Supplementary Information, Methods.

#### 2.4.2. LC-MS Raw Data Processing

Metabolite identifiers were harmonized through automated InChIKey-to-HMDB mapping against the Human Metabolome Database (HMDB version 5.0), followed by classification into endogenous and non-endogenous compounds (quality control in Supplementary Information, Methods). The data were subsequently transformed using centered log-ratio (CLR) transformation to account for the semi compositional nature of metabolomic measurements, and multivariate analyses were performed using CLR-transformed values. Biomarker discovery, feature selection, model optimization, and internal validation were performed exclusively in the training dataset, whereas the held-out test set was reserved for internal hold-out validation. For the targeted analyses, the selected metabolites and the seven composite biomarker indices were standardized as z-scores before statistical analysis.

#### 2.4.3. Integrated Systems Biology Pipeline

Rather than focusing solely on biomarker discovery, the analytical framework aimed to identify biologically meaningful metabolic networks linking metabolomic alterations to the dimensional clinical phenome via functional defined metabolic modules.

#### 2.4.4. Explorative Metabolomics Analysis

Within the discovery pipeline, the global metabolic structure was initially explored by principal component analysis (PCA) using CLR-transformed data. PCA was performed exclusively in the training dataset. The independent testing dataset (split ratio: test: 30%; training=70%) was subsequently projected into the PC space defined by the training dataset using the original loading matrix, without refitting or recalibrating the PCA model, thereby providing an external assessment of the robustness and generalizability of the derived metabolic modules.

Differential metabolite abundance between groups was evaluated on the non-CLR processed abundance data using two-sided Wilcoxon rank-sum tests, followed by Benjamini-Hochberg false discovery rate (FDR) correction and fold-change filtering (software and KEGG enrichment analysis in Supplementary Information, Methods).

#### 2.4.5. Machine Learning–Based Discovery and Validation of Metabolomic Biomarkers

Multivariate discrimination was performed on CLR-transformed data using partial least squares discriminant analysis (PLS-DA), orthogonal partial least squares discriminant analysis (OPLS-DA) and Random Forest (RF). Model performance of the PLS-DA and OPLS-DA models was evaluated using R²X, R²Y, predictive ability (Q²) statistics, permutation testing with permutation intercepts satisfying accepted criteria (R² intercept < 0.30 and Q² intercept < 0), and repeated nested cross-validation comprising five inner folds for model optimization and five outer folds for performance evaluation, repeated 10 times. RF models were likewise evaluated using nested cross-validation. Classification performance was quantified using balanced accuracy, sensitivity, specificity, cross-validated area under the receiver operating characteristic curve (CV-AUC), and the F1 score (validation criteria and calibration metrics in Supplementary Information, Methods).

#### 2.4.6. Feature Selection

The initial biomarker panel was identified through feature selection based on predictive performance, statistical robustness, and model stability. Candidate biomarkers were prioritized using an ensemble feature-selection strategy that integrated complementary statistical and machine-learning approaches, including Elastic Net penalized logistic regression, recursive feature elimination (RFE), bootstrap resampling (500 iterations), repeated stability selection, RF variable importance, Boruta feature selection, and sparse PLS-discriminant analysis (stability analyses in Supplementary Information, Methods). Each metabolite was assigned an evidence score according to the number of independent analytical methods supporting its selection, and only biologically annotated endogenous metabolites satisfying the predefined evidence criteria were retained in the final biomarker panels.

PLS-DA, OPLS-DA and RF models were used to validate this first selected metabolite panel (performance evaluation in Supplementary Information, Methods). A second biomarker panel was then derived using a systems biology framework that integrated biological relevance and representation of major metabolic pathways. To avoid multicollinearity, overfitting, and multiple testing burden while improving further model stability and interpretability, reduced biomarker panels were created by selecting top metabolites from the first biomarker panels. Given that the lipidomic biomarkers of MDD had already been characterized in the same dataset^18^, these variables were omitted from the present analyses to focus on non-lipid metabolic biomarkers. Using systems biology knowledge, the selected biomarkers were organized into functional metabolic modules. Modules were considered valid when a single principal component could be extracted from all constituent metabolites (KMO > 0.65, explained variance > 50%, and all factor loadings > 0.70), indicating coordinated regulation within a shared metabolic process.

#### 2.4.7. Regression Analysis

The final biomarker panel (n=20) together with the derived composite biomarker indices were subsequently analyzed in IBM SPSS Statistics version 30. Targeted statistical analyses included multivariate analysis of covariance (MANCOVA) to examine the differences in biomarkers between HC and FE-SDMD while adjusting for age, sex, BMI and MetS ranking. Linear discriminant function analysis (with leave one out cross validation) was used with diagnosis as dependent variables and the modules as explanatory variables. Correlation heatmaps were produced to display Pearsons’s product moment correlations between the module scores and demographic and clinical data.

Predictor selection for multiple regression models employed a multimethod strategy integrating manual regression analyses, automated variable-selection procedures, and biological knowledge. Candidate biomarkers were evaluated using overfitting-controlled regression, elastic net regularization, and best-subset regression. Final model selection was based not only on statistical performance and stability but also on biological plausibility and consistency with the metabolomics-based systems biology framework. The final set of metabolomic modules was subsequently entered into forced-entry multiple regression analyses to estimate their independent associations with the phenome dimensions. Because the primary objective was mechanistic inference rather than prediction, model evaluation emphasized robustness, stability, and biological interpretability instead of maximizing predictive performance. Model performance was evaluated using repeated k-fold cross-validation to estimate out-of-sample predictive ability and reduce overfitting (bootstrap stability, assumption checks, and performance indices in Supplementary Information, Methods).

#### 2.4.8. Individualized systems biology profiling

To capture biological heterogeneity across individuals, personalized biomarker contribution profiles were generated from the PLS-DA models. For each participant, the relative influence of the six metabolic module scores on the multivariate classification was quantified and visualized, providing a molecular fingerprint of the underlying metabolic disturbances. This strategy complements conventional case–control analyses by enabling characterization of subjects according to their individual biological signatures within a nomothetic systems biology framework.

## 3. Results

### 3.1. Sample and analytic overview

We studied 88 university students, 44 of which were healthy controls (HC) and 44 of which were patients with FE-SDMD. Supplementary Table 1 shows the demographic data of the subjects in the current study. Age, sex, and body mass index (BMI) were carried as covariates in every group-comparison model.

### 3.2. Explorative analysis

The untargeted plasma LC-QTOF-MS/MS platform returned 450 metabolites that passed quality control with no missing values. Initial univariate screening of the training dataset identified extensive metabolic differences between FE-SDMD patients and healthy controls (**Figure 1A**). Of the 450 detected metabolites, 306 reached nominal significance (raw *p* < 0.05), of which 298 remained significant after false discovery rate (FDR) correction (*q* < 0.05). Application of an additional fold-change threshold (FC > 1.5 or FC < 0.67) yielded 240 differentially abundant metabolites, comprising 119 upregulated and 121 downregulated metabolites in FE-SDMD. These metabolites constituted the initial candidate biomarker pool for subsequent multivariate analyses, feature selection, and construction of functional metabolic modules.

**Figure 1.**
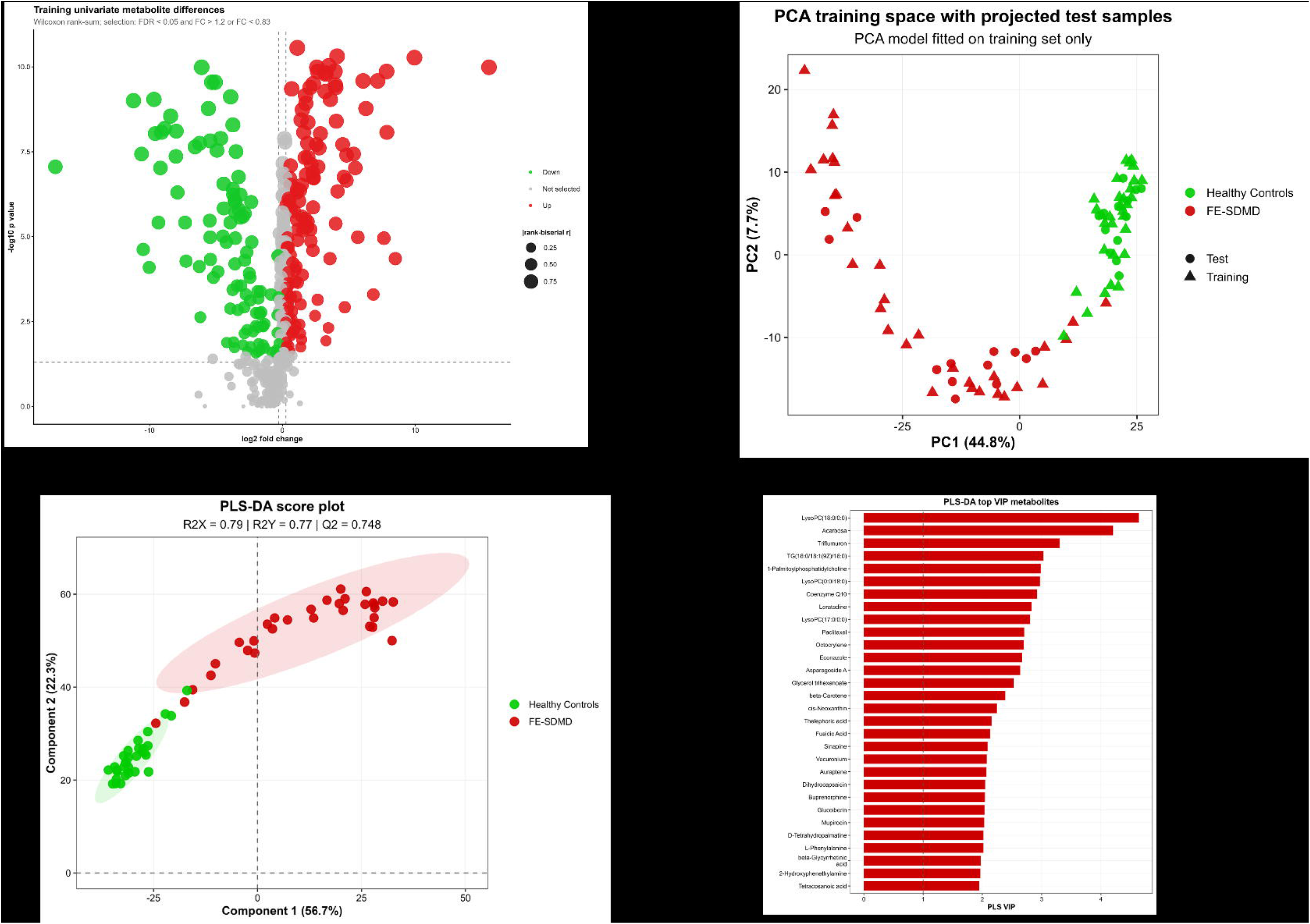
Volcano plot of differential metabolites in the training dataset. Differential metabolite abundance between patients with first-episode simple dysmood disorder (FE-SDMD) and healthy controls (HC) was assessed using the Wilcoxon rank-sum test (Figure 1A). The x-axis shows the log fold change (FE-SDMD versus HC), and the y-axis the −log (*p*) value. Metabolites meeting the selection criteria (FDR-adjusted *q* < 0.05 and fold change >1.5 or <0.67) are highlighted, with increased metabolites shown in red and decreased metabolites in green; gray symbols indicate metabolites that did not satisfy the selection criteria. Horizontal and vertical dashed lines denote the significance and fold-change thresholds, respectively. Of the 450 detected metabolites, 306 reached nominal significance (*p* < 0.05), 298 remained significant after FDR correction, and 240 fulfilled both the FDR and fold-change criteria, comprising 119 upregulated and 121 downregulated metabolites. The annotated metabolites represent those showing the largest and most significant changes and served as candidates for subsequent multivariate analyses, feature selection, and systems biology modeling. Figure 1B shows the principal component analysis (PCA) plot of the training dataset. The first PC (PC1) explained 44.8% of the total variance, whereas the second (PC2) accounted for an additional 7.7%. Figures 1C-1D show the results of PLS-DA. The shaded ellipses represent the 95% confidence regions for each group. The model showed good performance (R²X = 0.79, R²Y = 0.77, Q² = 0.748), with permutation testing (500 permutations) confirming that the observed discrimination was not attributable to chance (*p* < 0.002). Figure 1D shows the Variable Importance in Projection (VIP) ranking of the 30 highest-ranking metabolites.

The PCA model was exclusively fitted on the training set, while test samples were subsequently projected into the established PCA space without influencing model construction. The projected test samples closely followed the training-set group structure, supporting the out-of-sample stability and generalizability of the metabolic separation. PCA revealed a clear separation between FE-SDMD patients and healthy controls (Figure 1B). The first two principal components explained 52.5% of the total variance (PC1 = 44.8%, PC2 = 7.7%). Group discrimination was driven primarily by PC1, whereas PC2 captured additional within-group metabolic variation. The limited overlap between the two groups indicates that the global metabolomic profile of FE-SDMD differs substantially from that of healthy controls, supporting the presence of distinct underlying metabolic signatures.

Both PLS-DA and OPLS-DA showed strong discriminative ability with permutation testing (500 permutations) confirming that model performance exceeded chance expectations (both *p* < 0.002). Figure 1C show the results of PLS-DA (R²Y = 0.77; Q² = 0.748 and 0.751, respectively). Figure 1D shows that PLS-DA VIP analysis ranked metabolites according to their contribution to group discrimination. Supplementary **Figure 1A** shows the performance metrics with a balanced accuracy of 97.7%. A total of 107 metabolites exhibited VIP values >1 and were considered discriminative features.

KEGG over-representation analysis of the PLS panel pointed to lipid- and glycan-related biology (Supplementary Table 2).

### 3.3. Feature selection

Candidate biomarkers identified by VIP scores (>1) were not selected in isolation but integrated with complementary machine learning and regression-based feature-selection methods, including sparse PLS-DA, Elastic Net, recursive feature elimination, bootstrap selection, stability selection, and RF/Boruta (Supplementary Figures 2 and 3). We combined recursive feature elimination (5 metabolites) and RF/Boruta (82 confirmed metabolites) with Elastic Net (19 metabolites), bootstrap selection (4 metabolites), stability selection (26 metabolites) to form the multi-method evidence base for the first panel selection. A metabolite entered this first panel only if it was annotated as endogenous in HMDB, reached FDR < 0.05, and was supported by at least two independent selection methods.

This procedure yielded a 51-metabolite panel (**Supplementary Figure 4**). The OPLS-DA model based on the 51 selected metabolites (Supplementary Figure 4A) showed adequate performance (Supplementary Figure 4B) and maintained excellent discriminatory performance comparable to the full metabolite model, with an identical nested cross-validated AUC (Supplementary Figure 1B). Feature selection therefore achieved substantial dimensionality reduction with minimal loss of classification performance, supporting a more parsimonious metabolic signature.

The 51-metabolite panel was further curated by excluding lipids previously reported in our lipidomics study^18^, pharmaceuticals, environmental contaminants, and non-endogenous compounds without clear biological relevance to human metabolism. In contrast, dietary phytochemicals and their host–microbiome-derived metabolites (e.g., 6’’-*O*-acetyldaidzin) were retained because they may represent biologically meaningful markers of diet–microbiome interactions within the systems biology framework. Consequently, from this 51-metabolite panel, 20 metabolites were selected which were used for further analyses and to construct the composite indices that were carried into the confirmatory analyses below.

The OPLS-DA model based on the 20-biomarker panel showed excellent discrimination between FE-SDMD patients and healthy controls (Supplementary **Figure 5A/5B**), yielding R²X = 0.726, R²Y = 0.870, and Q² = 0.856. Nested cross-validation demonstrated adequate discriminative performance, yielding a balanced accuracy of 94.4%, sensitivity of 100%, specificity of 88.9%, an area under the receiver operating characteristic curve (AUC) of 0.998, and an F1 score of 94.7% (permutation, calibration, and Random Forest results in Supplementary Information, Results).

### 3.4. Group differences in the final 20-metabolite panel

**Supplementary Table 3** shows the results of a one-way MANCOVA comparing HC and FE-SDMD on the 20 selected metabolites with age, sex, and BMI as covariates. The multivariate between-group effect was large, indicating that the metabolite profile separated the two groups after adjustment (F(20, 64) = 31.01, p < 0.001). None of the covariates reached significance (age: F = 0.49, *p* = 0.961; sex: F = 1.31, *p* = 0.205; BMI: F = 0.78, *p* = 0.723). Supplementary Table 3 shows that all 20 metabolites differed between the groups at the univariate level (all *q* < 0.001). Relative to controls, FE-SDMD patients had significantly higher uridine-diphosphate, cholestone, acetylspermidine, PC(16:0/20:4), desoxycholic acid, isolithocholic acid, and deoxyciprinol, and lower CoQ10, cyclolycopene-1,5-diol, glycerol-trihexanoate, tocopherol, carotene, guanosine triphosphate, xanthylic acid, isobutyric acid, soya isoflavone, acetyldaidzin, cortisol, cortisone, and 3,4,5-trimethoxyphenylpropionic acid. The largest effects were for uridine-diphosphate (F(1, 83) = 235.25), glycerol-trihexanoate (F(1, 83) = 201.20), cyclolycopene-1,5-diol (F(1, 83) = 180.41), and CoQ10 (F(1, 83) = 159.45).

Linear discriminant analysis (LDA) identified a parsimonious panel of six metabolites— uridine 5′-diphosphate, 2,6-cyclolycopene-1,5-diol, coenzyme Q10, cholestenone, cortisone, and guanosine triphosphate—that optimally discriminated FE-SDMD patients from healthy controls. The resulting discriminant function was highly significant (_χ_² = 194.12, *df* = 6, *p* < 0.001) and showed an adequate canonical correlation (*r* = 0.951). Leave-one-out cross-validation yielded an overall classification accuracy of 98.9%, indicating robustness and generalizability of the selected biomarker panel.

Using prior systems biology knowledge, the 20 curated metabolites were grouped into six biologically meaningful metabolic modules representing coordinated metabolic processes. The composition of these modules is shown in **Table 1**. In addition, we computed a z unit based composite score as zOS–zAN, reflecting the balance of oxidative stress versus antioxidant defenses. A second MANCOVA evaluated these biologically defined composite indices (**Table 2**). A highly significant overall effect of diagnosis was observed (F(7,77) = 37.80, *p* < 0.001). Compared with controls, FE-SDMD was characterized by higher OS, OS/AN imbalance, gut– liver dysfunction, and mitochondrial dysfunction indices, together with lower antioxidant, HPA-axis, and dietary-biotransformation indices. LDA identified that 5 metabolic modules, namely zOS-zOA, mitochondrial stress, gut-liver dysfunction, diet and biotransformation, and HPA axis function optimally discriminated FE-SDMD patients from healthy controls (_χ_² = 104.59, df = 5, *p* < 0.001, canonical correlation = 0.845). Leave-one-out cross-validation yielded an overall classification accuracy of 90.9%.

**Table 1:** Functional modules of key metabolites with biological interpretation.

| **Module** | **Short name** | **Metabolites** | **Biological interpretation** |
|---|---|---|---|
| **1** | **Mitochondrial dysfunction (Mitochondrial bioenergetics)** | ↓ Coenzyme Q10,<br>↓ Guanosine triphosphate (GTP),<br>↓ Xanthylic acid (XMP),<br>↓ Glycerol trihexanoate | These metabolites reflect interconnected components of mitochondrial bioenergetics, including electron transport (Coenzyme Q10), guanine nucleotide metabolism (XMP and GTP), and lipid-derived energy supply (glycerol trihexanoate). Their coordinated reduction is consistent with impaired mitochondrial bioenergetics and diminished cellular energy production. |
| **2** | **Oxidative stress (Oxidative membrane remodeling)** | ↑ Cholestenone,<br>↑ N-acetylspermidine,<br>↑ PC(16:0/20:4) | Increased cholestenone, N-acetylspermidine, and arachidonate-containing PC(16:0/20:4) are consistent with increased sterol oxidation, altered polyamine metabolism, and oxidative membrane/lipid remodelling. Oxidative polyamine catabolism may additionally generate hydrogen peroxide and contribute to oxidative stress. |
| **3** | **Gut-liver dysfunction (Microbial bile acid metabolism)** | ↑ Deoxycholic acid,<br>↑ Isolithocholic acid,<br>↑ 27-Deoxy-5β-cyprinol,<br>↓ Isobutyric acid | Reduced isobutyric acid suggests altered microbial branched-chain amino-acid fermentation, whereas increased deoxycholic acid, isolithocholic acid, and 27-deoxy-5β-cyprinol indicate disturbances in microbial–hepatic bile-acid metabolism. Together, these changes suggest gut–liver metabolic dysregulation. |
| **4** | **DIET and phytochemical biotransformation** | ↓ Soyasapogenol B,<br>↓ 6''-O-Acetyldaidzin,<br>↓ 3-(3,4,5-Trimethoxyphenyl)propanoic acid | Reduced soy- and plant-derived phytochemicals may reflect lower dietary phytochemical exposure and/or altered intestinal absorption, gut microbial biotransformation, hepatic metabolism, and clearance, potentially reducing phytochemical-derived antioxidant and anti-inflammatory influences. |
| **5** | **HPA axis function (HPA-axis glucocorticoids)** | ↓ Cortisol,<br>↓ Cortisone | Cortisol and cortisone reflect systemic glucocorticoid activity and HPA-axis function, with additional roles in endocrine–immune regulation. Their coordinated reduction may indicate attenuated HPA-axis/adrenal glucocorticoid activity and diminished glucocorticoid-mediated immunoregulatory feedback. |
| **6** | **Antioxidant (Lipophilic antioxidant defense)** | ↓Coenzyme Q10,<br>↓ α-Tocopherol acetate,<br>↓ β-Carotene,<br>↓ 2,6-Cyclolycopene-1,5-diol<br>↓ Glycerol trihexanoate | Coenzyme Q10, α-tocopherol acetate, β-carotene, and cyclo-lycopene are lipophilic antioxidants that protect lipoproteins and cellular membranes against lipid peroxidation, while glycerol trihexanoate contributes to lipid-derived energy metabolism. Their coordinated reduction is consistent with diminished lipophilic antioxidant defenses and increased vulnerability to oxidative lipid damage. |

**Table 2.** Functional module composite score differences between first-episode simple dysmood disorder (FE-SDMD) patients and healthy controls (HC).

| **Index (z-score)** | **Mean HC** | **Mean FE-SDMD** | **F(1, 83)** | **q** |
|---|---|---|---|---|
| Antioxidant defenses (AN) | 0.786 (0.098) | –0.786 (0.098) | 124.54 | < 0.001 |
| Oxidative stress (OS) | –0.776 (0.100) | 0.776 (0.100) | 117.10 | < 0.001 |
| zOS–zAN | –0.834 (0.088) | 0.834 (0.088) | 174.64 | < 0.001 |
| Gut-liver dysfunction | –0.757 (0.103) | 0.757 (0.103) | 105.48 | < 0.001 |
| HPA axis function | 0.772 (0.110) | –0.772 (0.110) | 84.18 | < 0.001 |
| Diet and biotransformation | 0.719 (0.111) | –0.719 (0.111) | 81.75 | < 0.001 |
| Mitochondrial dysfunctions | –0.837 (0.088) | 0.837 (0.088) | 177.21 | < 0.001 |

*Note.* Contrasts derived from estimated marginal means (covariates: age, sex, BMI). HC = healthy
controls; FE-SDMD = first episode simple dysmood disorder; HPA = hypothalamic-pituitary-adrenal axis. All tests survived Benjamini-Hochberg FDR adjustment at q < 0.001.

### 3.5. Associations between metabolic modules and clinical phenotypes

**Figure 2** presents the zero-order Pearson correlations of the seven indices (six modules and one composite score) with demographic data, BMI, MetS ranking, depression severity, related phenotypes, and psychosocial adversity. There were no significant associations between any of those modules and age, sex, BMI or MetS ranking. The oxidative stress, zOS-zAN imbalance, gut–liver dysfunction, and mitochondrial dysfunction indices were positively associated with the overall phenome score, depressive symptom severity, physiosomatic symptoms, lifetime suicidal ideation, lifetime suicidal behaviors, and insomnia, whereas the antioxidant, HPA-axis, and dietary metabolism indices showed inverse associations with these clinical domains. Cognitive performance was positively associated with the antioxidant, HPA-axis, and dietary metabolism indices and inversely associated with the oxidative stress, zOS-zAN imbalance, gut–liver dysfunction, and mitochondrial dysfunction indices. Higher NLEs (interpersonal and academic relationships) and ACEs (particularly emotional neglect) were associated with higher oxidative stress, zOS-zAN imbalance, gut–liver dysfunction, and mitochondrial dysfunction indices, whereas the antioxidant, HPA-axis, and dietary metabolism indices showed inverse correlations. Collectively, these findings indicate that coordinated disturbances across the functional metabolic modules are closely linked to both clinical severity and psychosocial risk factors in FE-SDMD.

**Figure 2.**
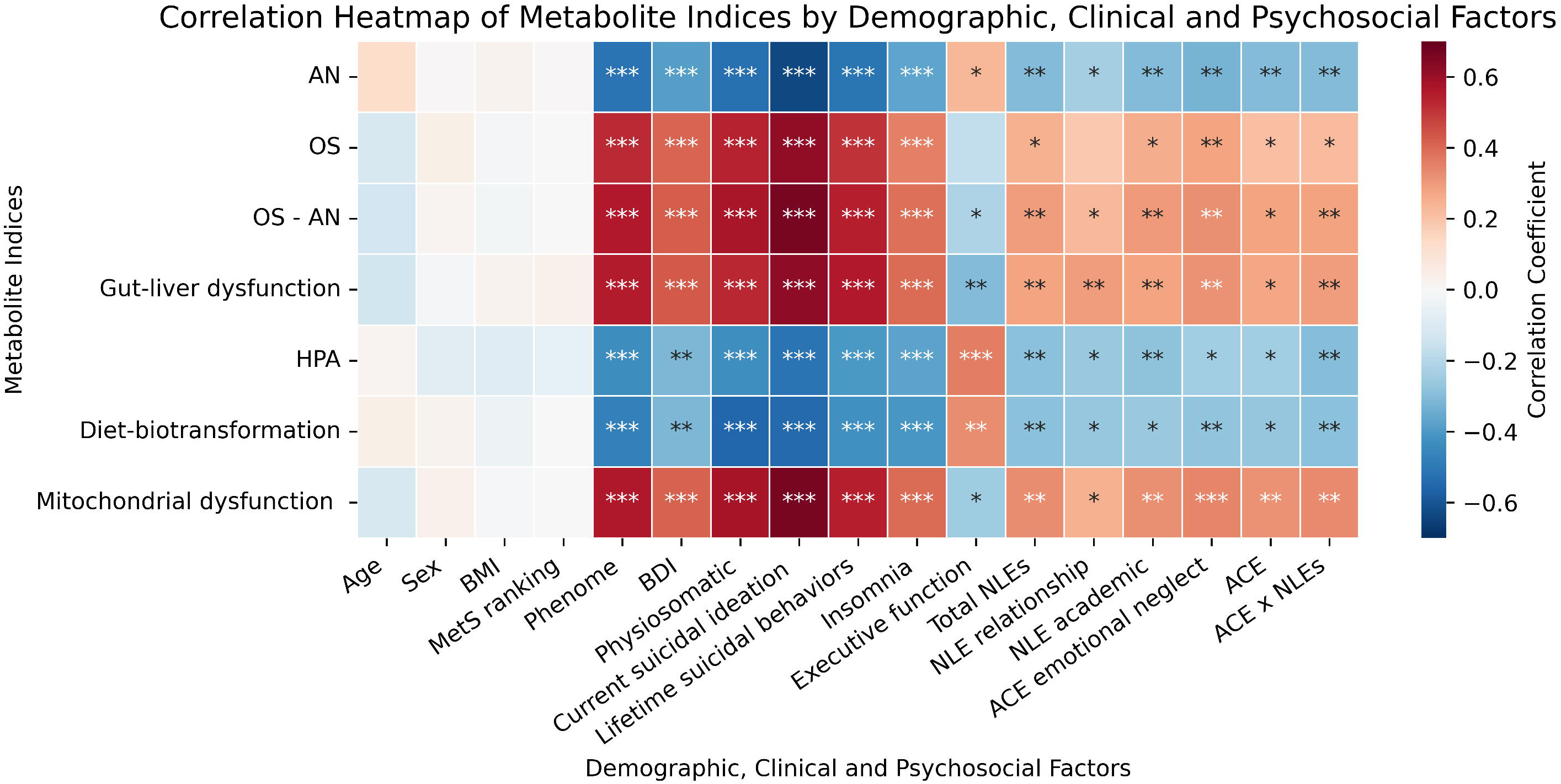
Correlations Between the Composite Biomarker Indices and Demographic factors, Clinical Phenotype, and Psychosocial Stressors. Cell values are Pearson correlations (*p < 0.05, **p < 0.01, ***p < 0.001). The color scale runs from negative (blue) to positive (red). AN = antioxidant capacity; OS = oxidative stress; OS-AN = zOS-zAN or oxidative stress/antioxidant ratio; HPA = hypothalamic-pituitary-adrenal axis; BMI = body mass index; SI = suicidal ideation; SB = suicidal behavior; BDI = pure Beck Depression Inventory; Physiosomatic = physiosomatic score; NLEs = Negative Live Events; ACE = adverse childhood experiences.

**Table 3** examines the combined contributions of psychosocial factors and composite metabolic modules to clinical phenotypes. All regression models were statistically significant and demonstrated substantial explanatory capacity across the evaluated domains. The overall phenome score was independently associated with NLE exposure, gut–liver dysfunction, and ACE-related emotional neglect, explaining 66% of the variance. The pure BDI-II score was significantly predicted (56.0% explained variance) by total NLEs, oxidative stress, and ACE emotional neglect, whereas physiosomatic symptoms were associated with mitochondrial stress, and total NLEs (R² = 0.41). Current suicidal ideation was independently related to gut–liver dysfunction, total NLEs, HPA-axis activity, and oxidative stress (R² = 0.43). Suicidal behaviors were explained (50% explained variance) by gut–liver dysfunction, total NLES, and total ACE exposure. Cognitive performance showed significant associations with HPA-axis activity and ACE exposure (R² = 0.20), whereas insomnia severity was predicted by total NLEs and dietary metabolism (R² = 0.37). Cross-validation yielded R² values ranging from 0.05 to 0.57, with the highest predictive performance observed for the overall phenome and suicidal behaviors. RMSE ranged from 0. 686 to 2.978 and MAE from 0.502 to 2.274, indicating variable prediction errors across the clinical outcomes.

**Table 3.** Regression of clinical phenotypes on combined psychosocial factors and composite biomarker indices.

| **Outcome (model fit)** | **Predictors** | **β** | **t** | **p** |
|---|---|---|---|---|
| ***Phenome*** |  |  |  |  |
| R² = 0.66, F(3, 84) = 54.67, p < 0.001 | Total NLEs | 0.54 | 7.90 | <0.001 |
| R² CV=0.572, RMSE=2.978, MAE=2.274 | Gut-liver dysfunction | 0.34 | 4.93 | <0.001 |
|  | ACE emotional neglect | 0.20 | 2.81 | 0.006 |
| ***Depression (BDI-II)*** |  |  |  |  |
| R² = 0.56, F(3, 84) = 35.16, p < 0.001 | Total NLEs | 0.553 | 7.054 | <0.001 |
| R² CV=0.378, RMSE=0.742, MAE=0.584 | Oxidative stress | 0.222 | 2.896 | 0.005 |
|  | ACE emotional neglect | 0.199 | 2.522 | 0.014 |
| ***Physiosomatic symptoms*** |  |  |  |  |
| R² = 0.41, F(2, 85) = 30.02, p < 0.001 | Mitochondrial dysfunction | 0.476 | 5.41 | <0.001 |
| R² CV=0.335, RMSE=0.770, MAE=0.582 | Total NLEs | 0.304 | 3.46 | <0.001 |
| ***Current suicidal ideation*** |  |  |  |  |
| R² = 0.43, F(4, 83) = 15.36, p < 0.001 | Gut-liver dysfunction | 0.54 | 3.41 | 0.001 |
| R² CV=0.393, RMSE=0.686, MAE=0.457 | Total NLEs | 0.36 | 4.16 | <0.001 |
|  | HPA axis function | 0.50 | 3.58 | <0.001 |
|  | Oxidative stress | 0.29 | 2.23 | 0.029 |
| ***Suicidal behaviors*** |  |  |  |  |
| R² = 0.50, F(3, 84) = 28.18, p < 0.001 | Gut-liver dysfunction | 0.40 | 4.94 | <0.001 |
| R² CV=0.396, RMSE=0.709, MAE=0.502 | Total NLEs | 0.31 | 3.62 | <0.001 |
|  | Total ACEs | 0.25 | 2.91 | 0.005 |
| ***Executive function*** |  |  |  |  |
| R² = 0.20, F(2, 85) = 10.38, p < 0.001 | HPA axis function | 0.29 | 2.89 | 0.005 |
| R² CV=0.053, RMSE=0.919, MAE=0.746 | Total ACE score | −0.27 | −2.70 | 0.008 |
| ***Insomnia (ISI)*** |  |  |  |  |
| R² = 0.37, F(2, 85) = 25.21, p < 0.001 | Total NLEs | 0.47 | 5.27 | <0.001 |
| R² CV=0.260, RMSE=0.808, MAE=0.667 | Diet and biotransformation | −0.27 | −3.05 | 0.003 |

*Note.* β = standardized coefficient; NLE = negative life event; ACE = adverse childhood experience; HPA = hypothalamic-pituitary-adrenal; HAM-D = Hamilton Depression Rating Scale; BDI = Beck Depression Inventory; ISI = Insomnia Severity Index. CV = cross-validated; RMSE = root mean square error; MAE = mean absolute error.

**Figure 3** shows score contribution plots of metabolic profiles in one healthy control (Figure 3A) and three FE-SDMD patients (Figure 3B, C, and D). In the Control, protective modules including the antioxidant defense, HPA-axis function, and diet-biotransformation contributed positively. Oxidative stress, gut–liver dysfunction, and mitochondrial dysfunction contributed negatively, and all with comparable magnitude. The three FE-SDMD patients exhibited distinct metabolic profiles. Patient 1 (B) showed pronounced abnormalities across all six modules, whereas Patient 2 (C) exhibited predominantly mitochondrial dysfunction and reduced antioxidant defenses. Patient 3 (D) showed mainly increased oxidative stress and gut– liver dysfunction, with relatively preserved other metabolic functions, illustrating marked interindividual heterogeneity.

**Figure 3.**
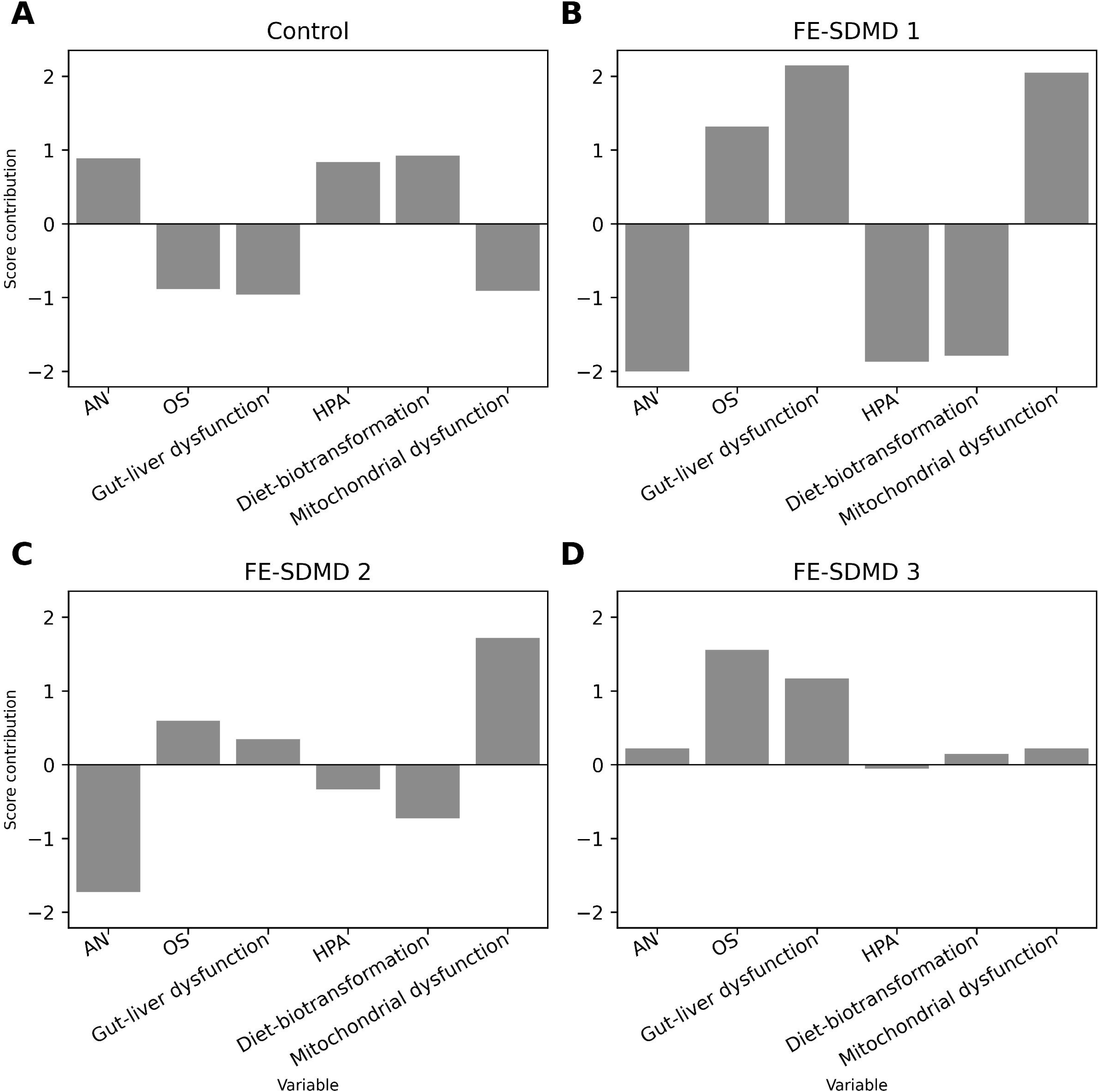
Personalized contribution plots illustrate varying contributions of metabolic indices in a healthy control and three outpatients with first-episode simple dysmood disorder (FE-SDMD). Panel A shows baseline metabolic contributions of a healthy control participant. Panel B, C, D illustrate different metabolic profiles across three FE-SDMD patients. AN = antioxidant capacity; OS = oxidative stress; HPA = hypothalamic-pituitary-adrenal axis

## 4. Discussion

### 4.1 Metabolic dysregulation in FE-SDMD

The first major finding of this study is that a 20-metabolite panel, organized into co-regulated functional metabolic modules, significantly distinguished the metabolic profile of FE-SDMD from that of controls. These modules captured mitochondrial stress and bioenergetic dysfunction, oxidative stress, gut–liver dysfunction, diet-related exposure and biotransformation, HPA-axis regulation, and antioxidant defenses. All modules differed significantly between FE-SDMD and HC, whereas none was significantly associated with age, sex, BMI, or subclinical MetS ranking. These findings indicate that FE-SDMD is characterized by a coordinated, systems-level metabolic signature that is not readily attributable to demographic factors, adiposity, or subclinical metabolic abnormalities.

Three of the six modules describe losses of protective systems. The lipophilic antioxidants CoQ10, α-tocopherol acetate, β-carotene, and cyclo-lycopene protect lipoproteins and cellular membranes against peroxidation. Their depletion is consistent with reduced reverse cholesterol transport^11^, and lower HDL antioxidant capacity^25^, vitamin E ^26^ and CoQ10 ^27^ levels in MDD.

The mitochondrial dysfunction module (lower CoQ10, GTP, XMP, and medium-chain triglycerides) may reflect impaired mitochondrial bioenergetics, encompassing reduced electron transport and guanine nucleotide metabolism alongside diminished lipid-derived energy supply. The findings are in line with lower mitochondrial respiration^28^, acetyl-L-carnitine and CoQ10 deficiencies in MDD^29^, and mitochondrial redox imbalances in MDD^20^.

Lower cortisol and cortisone levels in the HPA axis module contrast with the classical hypercortisolism in MDD, but blunted HPA-axis activity has been reported after childhood maltreatment^30,31^. In our work, the HPA-axis module was negatively related to ACE emotional neglect and to NLEs. Glucocorticoids are also endogenous immunoregulators that suppress proinflammatory cytokine production^32^, and negative feedback by cortisol is among the compensatory immune-regulatory system (CIRS)^33^. Lower plasma cortisol would therefore reduce this feedback. In the same cohort, FE-SDMD was marked by reduced CIRS activity without IRS activation ^34^ and a pre-proatherogenic lipid profile^11,34^. These findings support the concept that oxidative stress, and proatherogenic lipid alterations may occur across MDD phenotypes, whereas overt immune-inflammatory activation is more characteristic of severe and recurrent stages of MDD^19^.

Lower soy- and plant-derived dietary metabolites may indicate reduced dietary phytochemical and antioxidant exposure and/or altered gut microbial and host biotransformation, consistent with established associations between diet quality and depression^35^. In our study, reduced levels of diet-derived phytochemicals were additionally associated with greater insomnia severity. Although the mechanisms underlying this association remain unclear, previous reviews suggest that Mediterranean-style, antioxidant-rich, and polyphenol-rich dietary patterns may promote sleep quality, potentially through antioxidant, anti-inflammatory, metabolic, and gut– brain pathways^36,37^.

The remaining modules describe oxidative damage and gut dysbiosis. The OS module combines increased levels of cholestenone, n-acetylspermidine, and PC(16:00/20:4). An arachidonate-containing phosphatidylcholine increase is consistent with PLA2-mediated membrane remodeling and matches the PC(18:00/20:4) increase reported in inpatients^20^. Cholestenone is an oxidized sterol, and its presence may reflect oxidative stress^38^. N-acetylspermidine is a product of polyamine which generates hydrogen peroxide^39^. The gut-liver module (increased secondary bile acids and lower isobutyrate) implies altered microbial bile acid metabolism and protein fermentation, aligning with previous work^40^.

Uridine diphosphate (UDP), which was not assigned to any module, was significantly increased in MDD. UDP is an extracellular purinergic signaling molecule that activates microglial P2Y6 receptors and promotes phagocytosis^41^. In experimental models, increased UDP in the substantia nigra has also been linked to activation of mitochondrial ATP-sensitive potassium channels, attenuation of oxidative stress, and protection against mitochondrial dysfunction^42,43^. Thus, increased UDP in MDD could represent a compensatory purinergic response to mitochondrial and oxidative stress, although the relationship between circulating UDP and central purinergic signaling remains to be established.

### 4.2 Metabolomics, psychosocial stressors and the clinical phenome

The second major finding is that all metabolic modules were significantly associated with the key clinical severity indices. We observed that gut–liver dysfunction was the most consistent metabolic predictor of overall depression severity, current suicidal ideation, and lifetime suicidal behaviors, suggesting a particularly close relationship between gut–liver metabolic dysregulation and the suicidal phenome. Mitochondrial bioenergetic dysfunction was associated with greater physiosomatic symptom severity, whereas reduced HPA-axis activity was associated with more pronounced executive dysfunction. Previously, we showed that diacylglycerols and ceramides were strong predictors of overall depression severity and physiosomatic symptoms, whereas cholestenone and ceramides predicted suicidal behaviors^18^. Moreover, these lipid abnormalities, together with reduced lecithin–cholesterol acyltransferase (LCAT) activity and increased oxidative stress, explained substantial proportions of the variance in depression severity, physiosomatic symptoms, cognitive–affective symptoms, suicidal behaviors, insomnia, and anxiety^18^. Taken together, these findings indicate that distinct but interconnected gut–liver, mitochondrial, lipid-remodeling, oxidative, and neuroendocrine abnormalities map onto specific components of the depressive phenome.

The third major finding is that ACEs and NLEs, as well as their interactions, were associated with progressively greater abnormalities across the metabolic modules, suggesting that cumulative and interacting stressors are linked to broad systems-level metabolic dysregulation. Previously, we showed that ACEs and NLEs were strongly associated with lipidomics abnormalities, including ceramides, diacylglycerols, and cholestone, increased oxidative stress, and reduced antioxidant defenses^18–20^. These metabolic effects may constitute pathways through which ACEs and NLEs and their cumulative adversity effects become biologically embedded. Indeed, complex intersections between ACEs and NLEs are associated with the clinical phenome of depression, including suicidal behaviors and greater severity of depression, insomnia, and anxiety^44^. Taken together, these findings support a model whereby childhood and recent life stressors, and particularly their combined effects, are associated with coordinated NIMETOX abnormalities and more severe dimensional psychopathology.

### 4.3 Metabolomics and heterogeneity of MDD

Importantly, the functional metabolic abnormalities identified here in FE-SDMD outpatients substantially overlapped with those previously observed in acutely, more severely depressed inpatients, many of whom had recurrent MDD^20,45^. This convergence suggests that mitochondrial bioenergetic, lipid, gut–liver, and redox dysregulation is already present during the first episode of MDD and therefore cannot be explained solely by illness recurrence or chronicity. Moreover, because these abnormalities emerged in FE-SDMD in the absence of established metabolic syndrome, they are unlikely to represent merely secondary consequences of overt metabolic disease. Rather, the findings suggest that these interconnected metabolic and redox abnormalities constitute early components of MDD pathophysiology that may precede, and potentially contribute to, the progressive metabolic and immune abnormalities observed in more severe and recurrent illness.

We also showed that, although the metabolic modules significantly discriminated FE-SDMD from controls at the group level, individual metabolic profiles may provide additional clinically relevant information (Figure 3). FE-SDMD is metabolically heterogeneous: individual patients differ in the type, magnitude, and combination of abnormalities across mitochondrial bioenergetics, lipid and redox regulation, gut–liver function, HPA-axis activity, and diet/exposure-related metabolic signals. Consequently, the same clinical diagnosis may encompass markedly different metabolic configurations, while similar functional abnormalities may occur across patients with different symptom profiles. This heterogeneity may be obscured when biomarkers are interpreted as group means or diagnostic classifiers. A more informative precision-psychiatry approach would therefore construct an individualized metabolic profile for each patient, identifying the dominant dysfunctional modules and their relationships with specific phenome domains. Such personalized metabolic fingerprints could enable biologically informed stratification of MDD into outpatient FE-SDMD and inpatient MDD, and indeed mechanistically meaningful subgroups based on NIMETOX pathways^1,19,46^.

### 4.4 Limitations and future research

The first limitation is the cross-sectional design. Consequently, causal relationships, temporal changes in the metabolic modules, their long-term associations with symptom trajectories, and putative mediation or interaction effects cannot be established. Second, curation of the final 20-metabolite panel incorporated systems biology knowledge by the authors and therefore requires validation in independent cohorts. Third, some biologically informed modules are not completely independent; for example, the mitochondrial dysfunction and antioxidant modules share CoQ10. Rather than representing purely discrete pathways, however, such overlap also reflects the biological interconnectedness of mitochondrial bioenergetics, redox regulation, and antioxidant defenses. Fourth, because the untargeted metabolomics platform provides relative quantitative measurements, targeted assays using authentic standards are required to confirm metabolite identities and quantify absolute concentrations. Finally, in the absence of detailed dietary records, circulating phytochemical signals cannot be attributed specifically to differences in soy or other plants’ consumption or lower biotransformation.

### 4.5 Conclusions

In this study, untargeted LC-QTOF-MS/MS metabolomics revealed distinct metabolic profiles in young adults with FE-SDMD compared with healthy controls. These abnormalities converged into biologically informed, co-regulated modules reflecting lower antioxidant defenses, glucocorticoid activity, and diet-derived phytochemical exposure/biotransformation, and higher oxidative stress, gut–liver dysfunctions, and mitochondrial dysfunctions. These modules were differentially associated with symptom dimensions and psychosocial adversity, indicating that FE-SDMD is characterized not by a single metabolic abnormality but by heterogeneous and interconnected metabolic disturbances that map onto distinct components of the clinical phenome. The presence of mitochondrial, redox, lipid, and gut–liver abnormalities already during a first episode suggests that these alterations cannot be explained solely by illness chronicity or recurrence. Moreover, the marked interindividual variation in module profiles supports moving beyond diagnosis-level biomarker signatures toward personalized metabolic profiling capable of identifying the dominant biological disturbances in individual patients. Overall, these findings are consistent with the NIMETOX framework of MDD and support a systems-biology perspective integrating metabolic dysfunction, psychosocial adversity, and dimensional psychopathology. Given the preliminary and cross-sectional nature of the study, these observations should be regarded as hypothesis-generating and require confirmation in independent and longitudinal cohorts using targeted quantitative metabolomics.

## Data Availability

All data produced in the present study are available upon reasonable request to the authors after the authors have fully utilized the data

## Acknowledgements

VS was supported by the Scholarship from Graduate Affairs, Faculty of Medicine, Chulalongkorn University and the Ratchadaphisek Research Fund (Faculty of Medicine), MDCU (Batch#58), Chulalongkorn University, Thailand.

## CRediT authorship contribution statement

**Visuthsiri Sirivatanapa:** Conceptualization, Data Curation, Formal Analysis, Funding Acquisition, Investigation, Project Administration, Software, Visualization, Writing – original draft. **Pannipa Janta:** Data Curation, Methodology, Resources, Validation. **Asara Vasupanrajit:** Project Administration, Resources, Investigation. **Rossarin Tansawat:** Resources, Supervision. **Andre Carvalho**: Writing – Review & Editing, Validation. **Abbas F Almulla**: Formal Analysis, Writing – Review & Editing, Software. **Chavit Tunvirachaisakul:** Supervision, Resources, Writing – Review & Editing. **Michael Maes:** Conceptualization, Data Curation, Formal Analysis, Methodology, Project Administration, Supervision, Validation, Visualization, Writing – original draft, Writing – Review & Editing.

## Ethics approval

The research project (#0540/68) was approved by the Institutional Review Board of Chulalongkorn University’s institutional ethics board, Bangkok, Thailand, which follows the International Guideline for Human Research protection as required by the Declaration of Helsinki, The Belmont Report, CIOMS Guideline and International Conference on Harmonization in Good Clinical Practice (ICH-GCP). All participants gave informed consent.

