## Supplementary File for "Integrated Metabolomics Identifies Mitochondrial Dysfunction, Oxidative Membrane Remodeling, and Reduced Lipophilic Antioxidant Defenses in First-Episode Simple Dysmood Disorder"

**Supplementary Methods**

*Data Collection and Clinical Assessments (main text Section 2.2)*

Degree of anxiety severity was assessed by State-Trait Anxiety Inventory (STAI)^1^. Degree of depression was assessed by Hamilton Depression Scale (HAM-D) ^2^and Beck depression inventory-II (BDI-II)^3^. A “BDI” or “restricted cognitive-affective” score was calculated by summing all BDI-II items except appetite, tiredness, interest in sex, and sleep, thereby capturing the affective-cognitive aspects of depression. A z unit-based composite score of 3 HAM-D items (i.e., somatic anxiety, gastrointestinal somatic, and general somatic) and 2 BDI-II items (loss of energy and fatigue) were used to reflect the severity of physiosomatic symptoms. Insomnia and its severity were assessed using a Thai translated version ^4^ of the 7-item Insomnia Severity Index (ISI)^5^. The Thai translation of the ISI showed adequate internal consistency with Cronbach’s alpha of 0.90-0.91.

Suicidal behaviors were assessed using the Columbia-Suicide Severity Rating Scale (CSSRS) ^6^which was Thai translated byThe Columbia Lighthouse Project ^7^. The scale measures current and lifetime suicidal ideation (SI) and attempts (SA). Principal components (PC) were extracted from this scale to construct “suicide behavior” (SB)^8^. To describe shortly, PC SI lifetime was derived from the 10-item CSSR SI lifetime, and PC SI current from the 10-item CSSR SI within the past month. PC SA lifetime used CSSR SA lifetime items 1, 4, 6, 8, and 9, and PC SA current used CSSR SA current items 1, 2, 4, 6, 8, and 9^8^. PC SB lifetime = PC SI lifetime + PC SA lifetime; PC SB current = PC SI current + PC SA current; and total SB = PC SB lifetime + PC SB current^8^.

The Thai translation ^9^ of the Adverse Childhood Experiences (ACE) Questionnaire ^10^ was used to measure domains of abuse (e.g., emotional, physical, and sexual), showing internal consistency of 0.79, and neglect (e.g., physical and emotional), showing internal consistency of 0.82. ACE load was computed by the total scale score^11^. Negative life events (NLEs) were measured by the Negative Event Scale ^12^ which was Thai-translated^13^. The scale measures both interpersonal and non-interpersonal NLEs. In the current study we used three sum scores: total NLE (all items), relationship-linked items, and academic items^14^.

*Metabolic Syndrome Criteria (main text Section 2.2)*

The criteria include central obesity (waist circumference > 90 cm in men, > 80 cm in women), triglycerides ≥ 150 mg/dL and low high-density lipoprotein (< 40 mg/dL in men and < 50 mg/dL in women), blood pressure ≥ 130/85 mmHg, and fasting glucose ≥100 mg/dL^15^. Individuals that satisfied at least three of these criteria were excluded from the study.

*Assays (main text Section 2.3)*

Plasma samples were prepared following the previously published methods ^16,17^ and subsequently analyzed using ultra-high-performance liquid chromatography coupled with quadrupole time-of-flight tandem mass spectrometry (UHPLC–QTOF–MS/MS). In brief, 200 µL of plasma was extracted with a cold methanol-ethanol mixture (1:1, v/v)^18^, vortex-mixed, sonicated, and centrifuged at 14,000 rpm for 20 min at 0 °C (Centrifuge 5810 R, Eppendorf, Hamburg, Germany). The resulting supernatant was dried using a vacuum centrifuge (2000 rpm; SCANVAC, Labogene ApS, Lynge, Denmark), reconstituted in methanol-water (4:1, v/v), filtered through a 0.22 *µ*m PTFE membrane, and transferred into HPLC vials. The prepared samples were stored at –80°C until analysis. A pooled quality control (QC) sample was prepared by combining 200 *µ*L aliquots from each plasma sample and processed using the same extraction procedures as the study samples.

Untargeted metabolomic analysis was performed using UHPLC–QTOF–MS/MS consisting of a Dionex Ultimate 3000 UHPLC system (Thermo Fisher Scientific) coupled to a Bruker impact II QTOF MS (Bruker Daltonics, Bremen, Germany). Chromatographic separation was done using a Halo 120 C18 column (100 mm × 2.1 mm, 2.7 µm) maintained at 40 °C with the autosampler temperature set at 7 °C. Gradient elution was performed at a flow rate of 0.3 mL/min with an injection volume of 1 µL as follows: 0–15 min, 0–99% B; 15–20 min, 99% B; and 20–25 min, re-equilibration to 1% B^19^.

Mass detection used a scanning m/z 50–1200, with collision energies of 20.0 eV (positive mode) and 10.0 eV (negative mode), both were electrospray ionization modes^19^. Nitrogen was used as both the nebulizing and collision gas, with a dry gas temperature of 250 °C, a dry gas flow rate of 8.0 L/min, and a capillary voltage of 3800 V. pooled QC samples were analyzed after every 10 study samples to monitor analytical stability throughout the analytical batch.

Raw data were converted to .abf file before being processed in MS-DIAL v5.5.241113 for peak detection, spectral deconvolution, retention-time alignment, and metabolite identification. Metabolite annotation was performed by matching MS/MS spectra against the PubChem, the Human Metabolome (HMDB), and MassBank database. Metabolites were retained only if they exhibited a fold change ≥ 1.5 relative to the blank sample and a signal-to-noise (S/N) ratio ≥ 20.

*Classical Statistical Analyses (main text Section 2.4.1)*

Contingency table analyses with χ² tests were used to examine associations among categorical variables.

Sample size estimation was based on previous NIMETOX studies by Maes and colleagues, demonstrating that multivariable regression models incorporating metabolic biomarkers typically explain more than 25% of the variance in the clinical phenome (R² > 0.25). An a priori power analysis for the primary analyses, namely multiple regression models predicting the overall severity of depression score (the primary outcome measure), was performed using G*Power 3.1.9.4. Assuming an effect size corresponding to R² = 0.25 (f² = 0.33), a two-sided α of 0.05, a statistical power of 0.80, and a maximum of seven predictors, the required minimum sample size was 51 participants. To facilitate model development and independent validation, the final sample size was increased to permit division into training and testing datasets.

*LC-MS Raw Data Processing (main text Section 2.4.2)*

Metabolite abundances underwent quality-control assessment before missing values (if any) were imputed using the half-minimum approach. Nevertheless, the data used in the current study was free of any missing value.

*Integrated Systems Biology Pipeline (main text Section 2.4.3)*

Statistical and machine learning analyses were performed using **R** (version 4.6.1), **Statistica 14.0,** and **SPSS version 30.**

*Explorative Metabolomics Analysis (main text Section 2.4.4)*

Volcano plots, heatmaps, and hierarchical clustering analyses were generated using R packages and OmicStudio, with heatmaps constructed using the R package heatmap. Functional interpretation of the final biomarker panels was performed using KEGG pathway enrichment analyses. These analyses were intended solely to support biological interpretation of the validated biomarker panels and were not used as additional feature-selection procedures.

*Machine Learning–Based Discovery and Validation (main text Section 2.4.5)*

These approaches were considered supportive when they achieved predefined validation criteria, including positive CV Q² where applicable, statistically significant permutation testing, acceptable permutation intercepts, reproducible feature-selection frequencies across repeated resampling, a CV-AUC ≥ 0.75, a CV-balanced accuracy ≥ 0.70, and F1 > 0.75. The robustness of the machine-learning models was further evaluated using repeated bootstrap resampling. Model calibration was assessed using calibration curves, calibration intercepts, calibration slopes, and the Brier score. Good calibration was defined by a calibration intercept close to zero, a calibration slope close to one, and a low Brier score, indicating close agreement between predicted probabilities and observed outcomes.

*Feature Selection (main text Section 2.4.6)*

Bootstrap and stability analyses quantified the reproducibility of feature selection across repeated resampling, whereas sparse PLS-discriminant analysis provided an additional assessment of multivariate feature stability. In addition, the predictive performance of this first biomarker panel was evaluated using repeated nested cross-validation consisting of ten repetitions of five-fold outer cross-validation with internal hyperparameter optimization.

Because the primary objective was to construct and internally validate biologically meaningful metabolic modules rather than optimise diagnostic classification, nested cross-validation was used to assess the discriminative stability of the selected metabolite panels, while module validity was evaluated primarily through biological coherence, principal component structure, and associations with dimensional clinical phenotypes.

*Regression Analysis (main text Section 2.4.7)*

**Combining complementary statistical approaches with mechanistic biological interpretation facilitated the construction of robust, parsimonious, and biologically meaningful models while reducing the risk of overfitting.** **Model assumptions were verified by assessing residual normality with probability–probability (P–P) plots, multicollinearity with variance inflation factors (VIF) and tolerance values, and homoscedasticity using White's and the modified Breusch–Pagan tests.** Predictive performance was summarized using the cross-validated coefficient of determination (R²), root mean square error (RMSE), mean absolute error (MAE), and additional model-specific performance indices where appropriate. **Partial regression plots were constructed to illustrate the independent relationships between individual biomarkers and the phenome after adjustment for all remaining covariates.**

**Internal model stability was examined by bootstrap resampling (1,000 iterations). Regression models were summarized using standardized regression coefficients (β), t statistics, exact p-values, and coefficients of determination (R²).**

**Supplementary Results**

*KEGG Pathway Enrichment (main text Section 3.2)*

The most strongly enriched pathways were aldosterone-regulated sodium reabsorption, Ras signaling, ether-lipid metabolism, and several glycosphingolipid and O-glycan biosynthesis routes, along with complement and coagulation cascades, type II diabetes mellitus, and mucin-type o-glycan biosynthesis. All pathways showed p < 0.005.

*Sparse PLS-DA (main text Section 3.2)*

Supplementary Figure 2 shows the results of sparse PLS-DA indicating adequate discrimination between FE-SDMD patients and healthy controls, with a cross-validated AUC of 0.999 and a balanced accuracy of 0.977. Permutation testing confirmed that the observed classification performance was highly significant (500 permutations, p = 0.00498), indicating that the model was not overfitted. The sparse PLS-DA model identified a parsimonious subset of discriminatory metabolites, and selection-frequency analysis demonstrated that several metabolites were consistently retained across repeated model fitting.

*Feature Selection (main text Section 3.3)*

This strategy generated a consensus-based, statistically robust and biologically informed candidate biomarker panel for subsequent systems biology analyses. For example, in Supplementary Figure 3, feature importance was evaluated using RFE and the Boruta algorithm. The substantial overlap between RFE and Boruta rankings indicates that these metabolites are robust and reproducible discriminators of FE-SDMD.

Hierarchical clustering of the selected biomarkers revealed two distinct metabolic signatures that clearly differentiated FE-SDMD patients from healthy controls (Supplementary Figure 4A). The latter figure shows that many metabolites exhibited marked reductions in FE-SDMD, including α-tocopherol acetate, coenzyme Q10, glycerol trihexanoate, cortisone, cortisol, fasciculol C, sclareol, tetracosanoic acid, auraptene, and 6''-O-acetyldaidzin, whereas β-glycyrrhetinic acid, TG(16:0/18:1(9Z)/16:0), thelephoric acid, uridine 5′-diphosphate, and 1-methyl-2-nonyl-4(1H)-quinolinone were significantly increased. Principal component analysis of the fixed biomarker panel confirmed this separation, with PC1 explaining 64.3% and PC2 7.5% of the total variance (Supplementary Figure 4B). Importantly, both the training and independent testing samples clustered according to diagnostic status, indicating that the selected biomarker panel was robust, reproducible, and generalizable across datasets.

*Final 20-Metabolite Panel (main text Section 3.3)*

Permutation testing confirmed that the model was highly significant and not overfitted, with R²Y and Q² permutation p values both <0.01 and acceptable intercepts (R²Y = 0.082; Q² = –0.214). Calibration analysis performed in the independent test set demonstrated close agreement between predicted probabilities and observed outcomes, indicating good model calibration. Variable importance analysis (Supplementary Figure 5C) identified glycerol trihexanoate, uridine 5′-diphosphate, coenzyme Q10, 2,6-cyclolycopene-1,5-diol, N8-acetylspermidine, deoxycholic acid, and 6''-O-acetyldaidzin as the principal contributors to class discrimination.

RF analysis further confirmed the robustness of the 20-panel biomarker panel (Supplementary Figure 5D). Variable importance analysis identified cortisone, uridine 5′-diphosphate, 6''-O-acetyldaidzin, glycerol trihexanoate, 2,6-cyclolycopene-1,5-diol, coenzyme Q10, α-tocopherol acetate, and 3-(3,4,5-trimethoxyphenyl)propanoic acid as the strongest contributors to classification. Bootstrap stability analysis demonstrated consistently high selection frequencies for the majority of these biomarkers, with several metabolites being retained in nearly all bootstrap resamples. The concordance between variable importance and bootstrap stability indicates that the selected biomarkers are reproducible and not driven by sampling variability, thereby supporting their inclusion in the final consensus biomarker panel.

**Supplementary Tables**

**Supplementary Table 1.** *Participants’ Demographic Data, Clinical Characteristics, and Biomarkers*

| **Variable** | **Controls (n = 44)**  **Mean (SD)** | **FE-SDMD (n = 44)**  **Mean (SD)** | **p** |
| --- | --- | --- | --- |
| Age (years) | 23.477 (3.181) | 21.933 (3.298) | 0.028 |
| *Sex, n (%)* |  |  |  |
| Female | 37 (84.1%) | 38 (86.4%) | 0.764 |
| Male | 7 (15.9%) | 6 (13.6%) |  |
| BMI (kg/m²) | 22.331 (3.574) | 21.705 (4.899) | 0.495 |
| Years of education | 16.568 (1.485) | 15.523 (2.799) | 0.031 |
| *Current smoking, n (%)* |  |  |  |
| No | 43 (97.7%) | 39 (88.6%) | 0.091 |
| Yes | 1 (2.3%) | 5 (11.4%) |  |
| *Metabolic syndrome, n (%)* |  |  |  |
| No | 44 (100.0%) | 44 (100.0%) | 1 |
| MetS ranking score | 0.409 (0.693) | 0.409 (0.658) | 1 |
| OSOD | 0.636 (0.685) | 3.023 (1.355) | <0.001 |
| HAM-D physiosomatic symptoms | 0.73 (0.79) | 4.21 (1.97) | <0.001 |
| Cognitive-affective BDI-II | 3.26 (6.491) | 12.82 (7.25) | <0.001 |
| BDI-II | 6.773 (6.491) | 24.136 (12.233) | <0.001 |
| ISI | 5.591 (4.304) | 12.341 (6.416) | <0.001 |
| Current SI | 0 (0) | 1.864 (1.936) | <0.001 |
| Suicidal behavior score^$^ | −0.932 (0.197) | 0.505 (0.921) | <0.001 |
| Total STAI state score | 46.59 (14.14) | 59.25 (8.08) | <0.001 |
| Total NLE score | 42.20 (28.11) | 77.77 (27.72) | <0.001 |
| Total ACE score | 17.32 (6.49) | 24.41 (8.40) | <0.001 |

*Note.* Group differences were tested with the t-test and, for categorical variables, the χ² test. Values are mean (SD) unless otherwise stated. ^$^ z-scores are presented as Mean (SE). FE-SDMD = first-episode simple dysmood disorder; BMI = body mass index; OSOD =overall severity of depression; HAM-D = Hamilton Depression Rating Scale (somatic anxiety + gastrointestinal + general somatic items); BDI-II = Beck Depression Inventory-II; ISI = Insomnia Severity Index; SI = suicide ideation; STAI = State-Trait Anxiety Inventory; NLE = Negative Life Event; ACE = adverse childhood experience.

**Supplementary Table 2.** *Top KEGG pathways (mummichog over-representation)*

| **KEGG pathway** | **ID** | **Enrichment p** |
| --- | --- | --- |
| Aldosterone-regulated sodium reabsorption | hsa04960 | 0.0002 |
| Ras signaling pathway | hsa04014 | 0.0003 |
| Ether-lipid metabolism | hsa00565 | 0.0007 |
| Glycosphingolipid biosynthesis (globo/isoglobo) | hsa00603 | 0.0012 |
| Glycosphingolipid biosynthesis (lacto/neolacto) | hsa00601 | 0.0016 |
| Prostate cancer / choline-related signaling | hsa05215 | 0.0015 |
| Complement and coagulation cascades | hsa04610 | 0.0016 |
| Type II diabetes mellitus | hsa04930 | 0.0020 |
| Mucin-type O-glycan biosynthesis | hsa00512 | 0.0024 |

*Note.* Fold-change threshold: FC > 1.2 or < 0.83. Enrichment p = mummichog/KEGG over-representation score (lower = stronger); foreground = 33 mapped compounds from the PLS panel.

**Supplementary Table 3.** *Covariate-adjusted estimated marginal means and between-subjects effects for 20 plasma metabolites by diagnostic group*

| **Metabolite (z-scores)** | **Mean HC (SE)** | **Mean FE-SDMD (SE)** | **F(1, 83)** | **q** |
| --- | --- | --- | --- | --- |
| Uridine-diphosphate | −0.87 (0.079) | 0.87 (0.079) | 235.25 | < 0.001 |
| CoQ10 | 0.81 (0.089) | −0.81 (0.089) | 159.45 | < 0.001 |
| Guanosine triphosphate | 0.46 (0.139) | −0.46 (0.139) | 21.17 | < 0.001 |
| Xanthylic acid | 0.61 (0.125) | −0.61 (0.125) | 46.15 | < 0.001 |
| Glycerol-trihexanoate | 0.84 (0.082) | −0.84 (0.082) | 201.20 | < 0.001 |
| Tocopherol | 0.65 (0.120) | −0.65 (0.120) | 57.15 | < 0.001 |
| Carotene | 0.62 (0.125) | −0.62 (0.125) | 47.39 | < 0.001 |
| Cyclolycopene-1,5-diol | 0.82 (0.085) | −0.82 (0.085) | 180.41 | < 0.001 |
| Cholestone | −0.59 (0.127) | 0.59 (0.127) | 42.52 | < 0.001 |
| Acetylspermidine | −0.73 (0.107) | 0.73 (0.107) | 88.98 | < 0.001 |
| PC(16:0/20:4) | −0.59 (0.126) | 0.59 (0.126) | 43.27 | < 0.001 |
| Deoxycholic acid | −0.61 (0.118) | 0.61 (0.118) | 51.85 | < 0.001 |
| Isolithocholic acid | −0.58 (0.124) | 0.58 (0.124) | 41.83 | < 0.001 |
| Deoxyciprinol | −0.49 (0.137) | 0.49 (0.137) | 25.19 | < 0.001 |
| Isobutyric acid | 0.55 (0.131) | −0.55 (0.131) | 35.00 | < 0.001 |
| Cortisol | 0.65 (0.118) | −0.65 (0.118) | 58.21 | < 0.001 |
| Cortisone | 0.74 (0.108) | −0.74 (0.108) | 90.65 | < 0.001 |
| Soya isoflavone | 0.64 (0.121) | −0.64 (0.121) | 54.46 | < 0.001 |
| Acetyldaidzin | 0.68 (0.116) | −0.68 (0.116) | 67.26 | < 0.001 |
| 3,4,5-Trimethoxyphenylpropionic acid | 0.67 (0.116) | −0.67 (0.116) | 65.59 | < 0.001 |

*Note.* Means are covariate-adjusted estimated marginal means on a z-score scale (covariates: age, sex, BMI). HC = healthy controls; FE-SDMD = first-episode simple dysmood disorder. All comparisons remained significant after Benjamini-Hochberg false discovery rate adjustment.

**Supplementary Figures**

Supplementary Figure 1

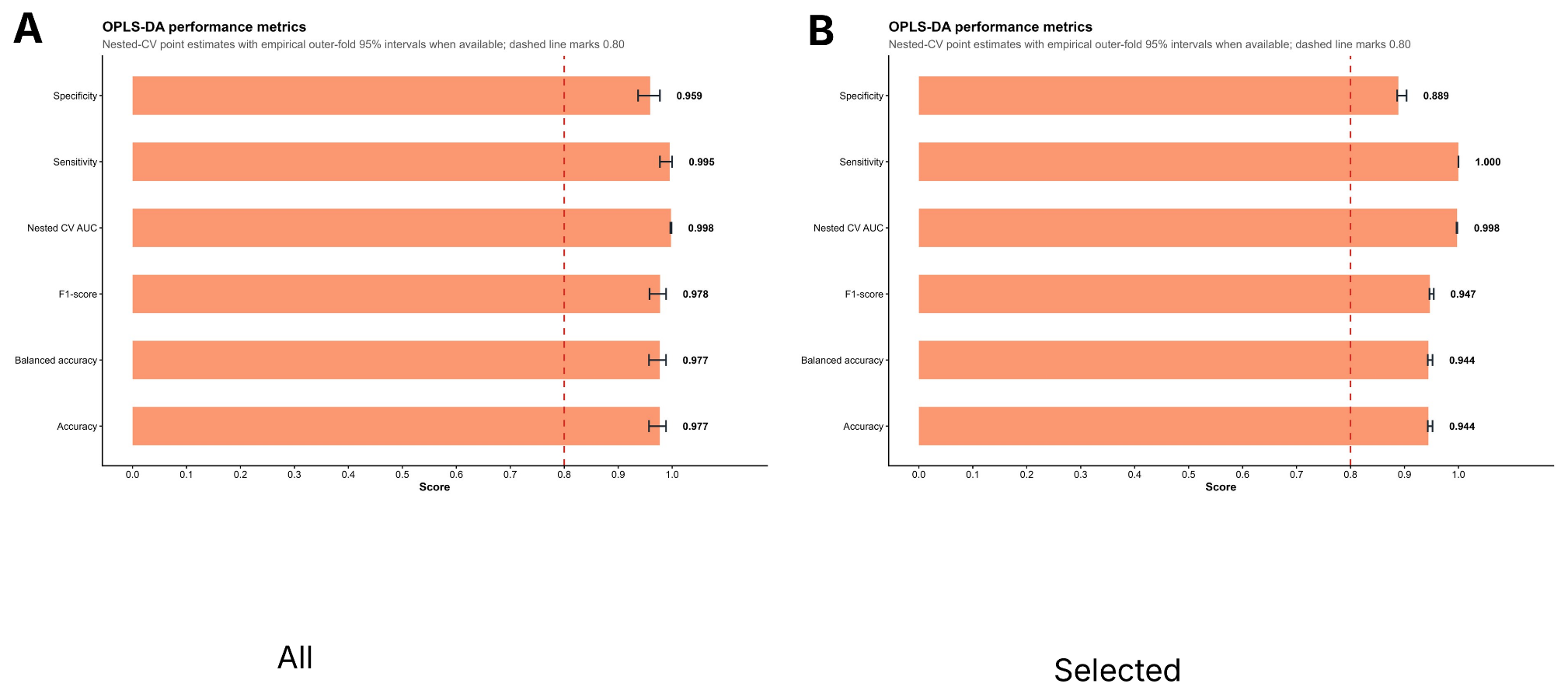

Supplementary Figure 2

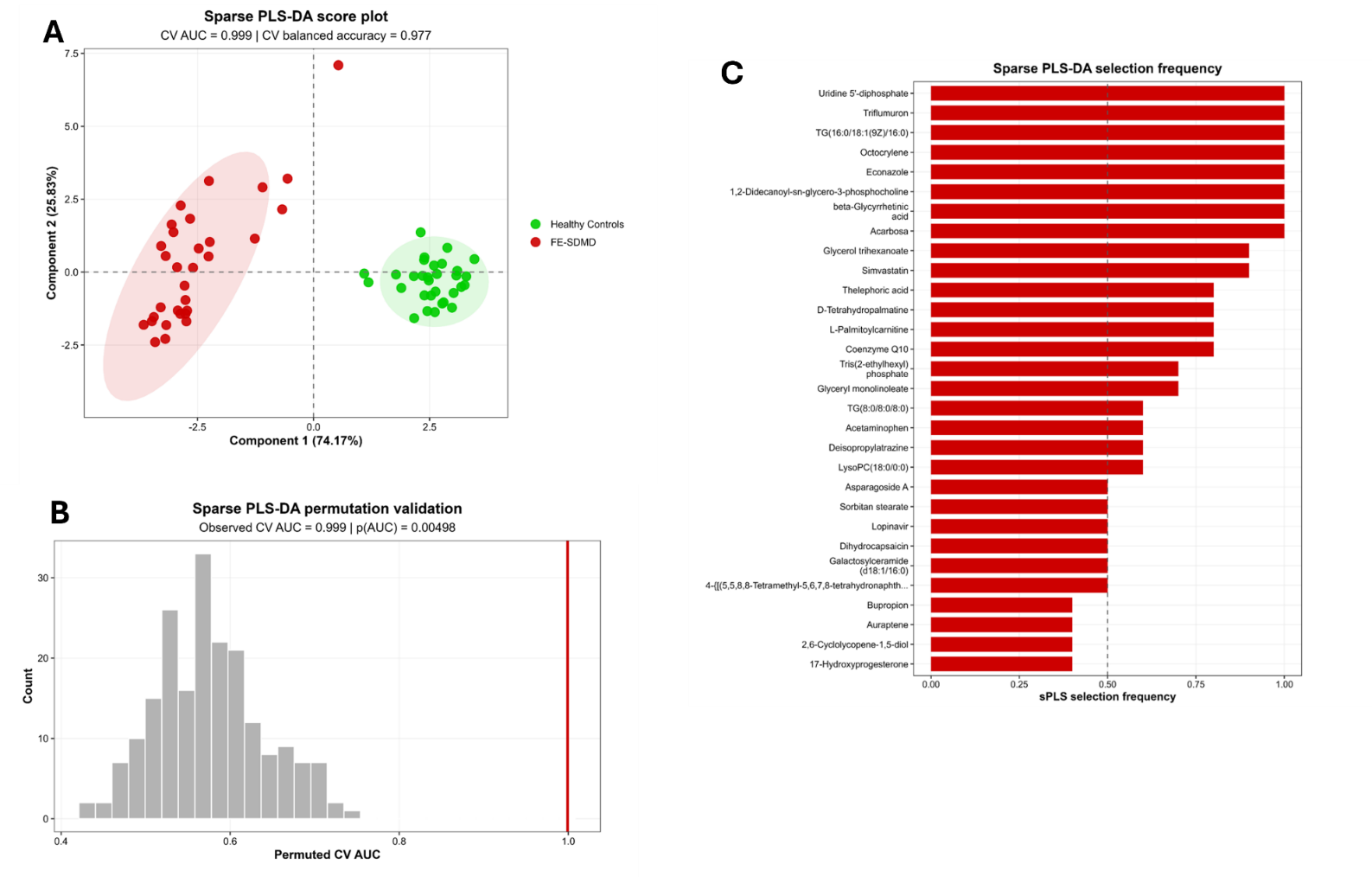

Supplementary Figure 3
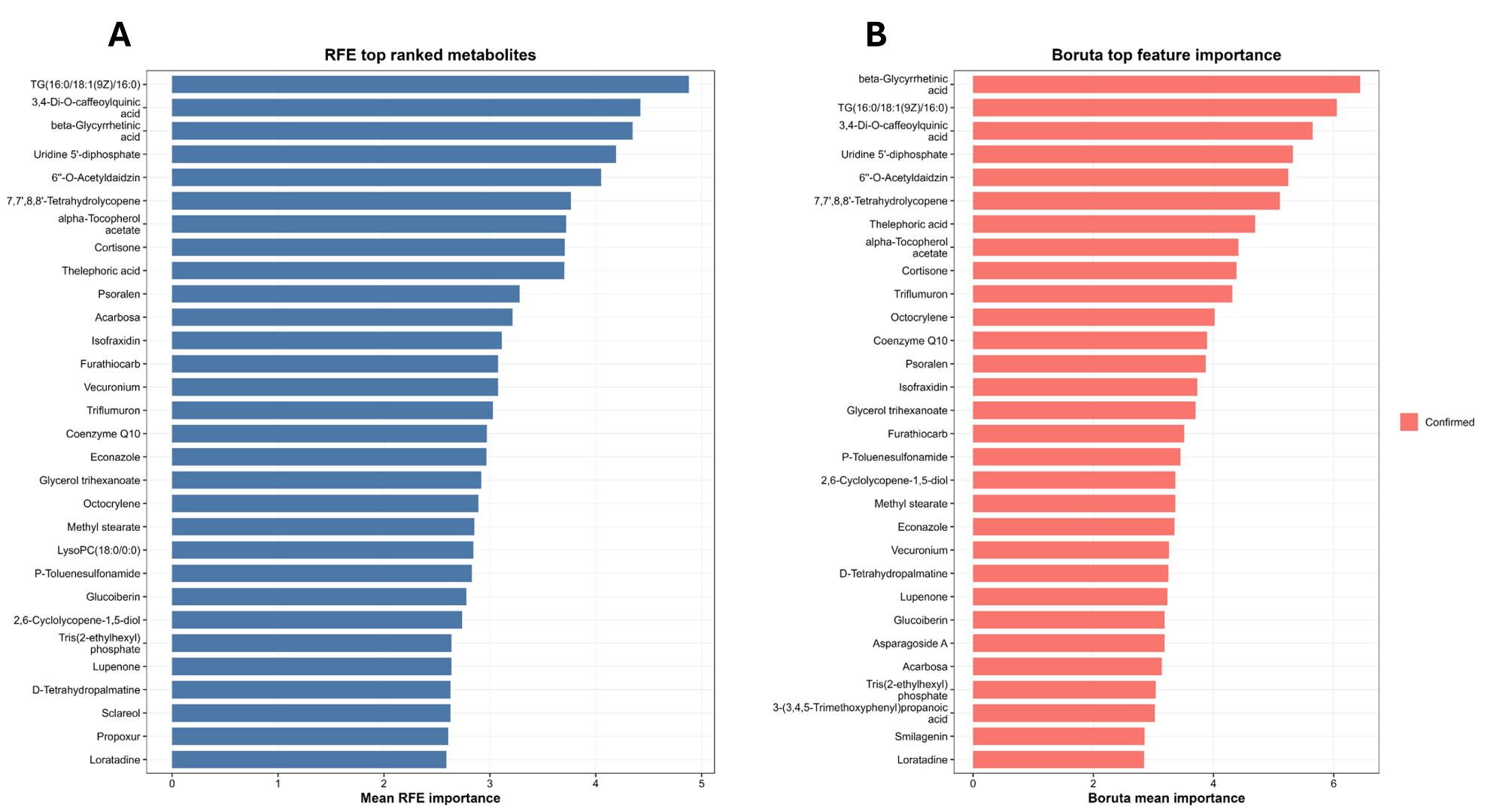

Supplementary Figure 4

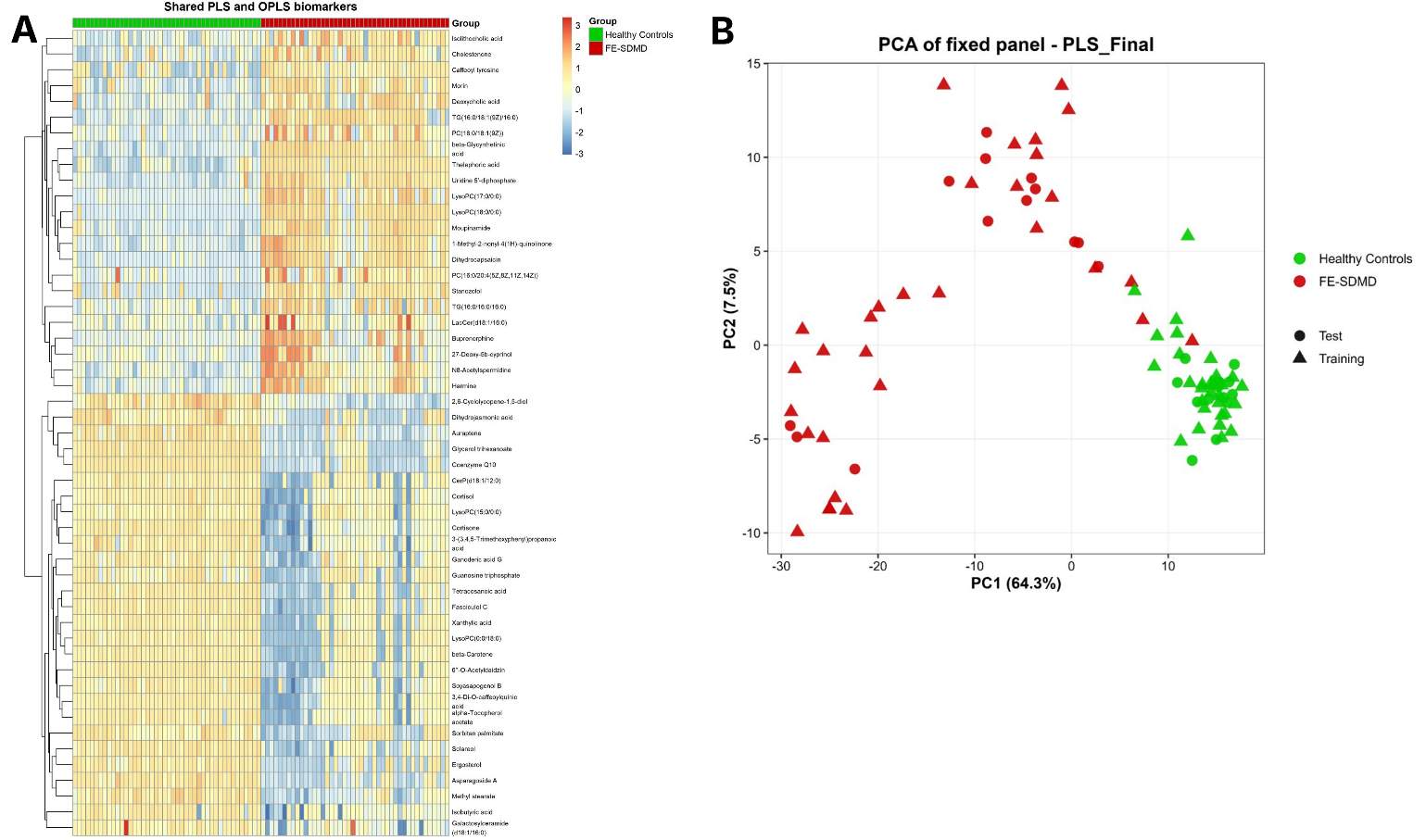

Supplementary Figure 5

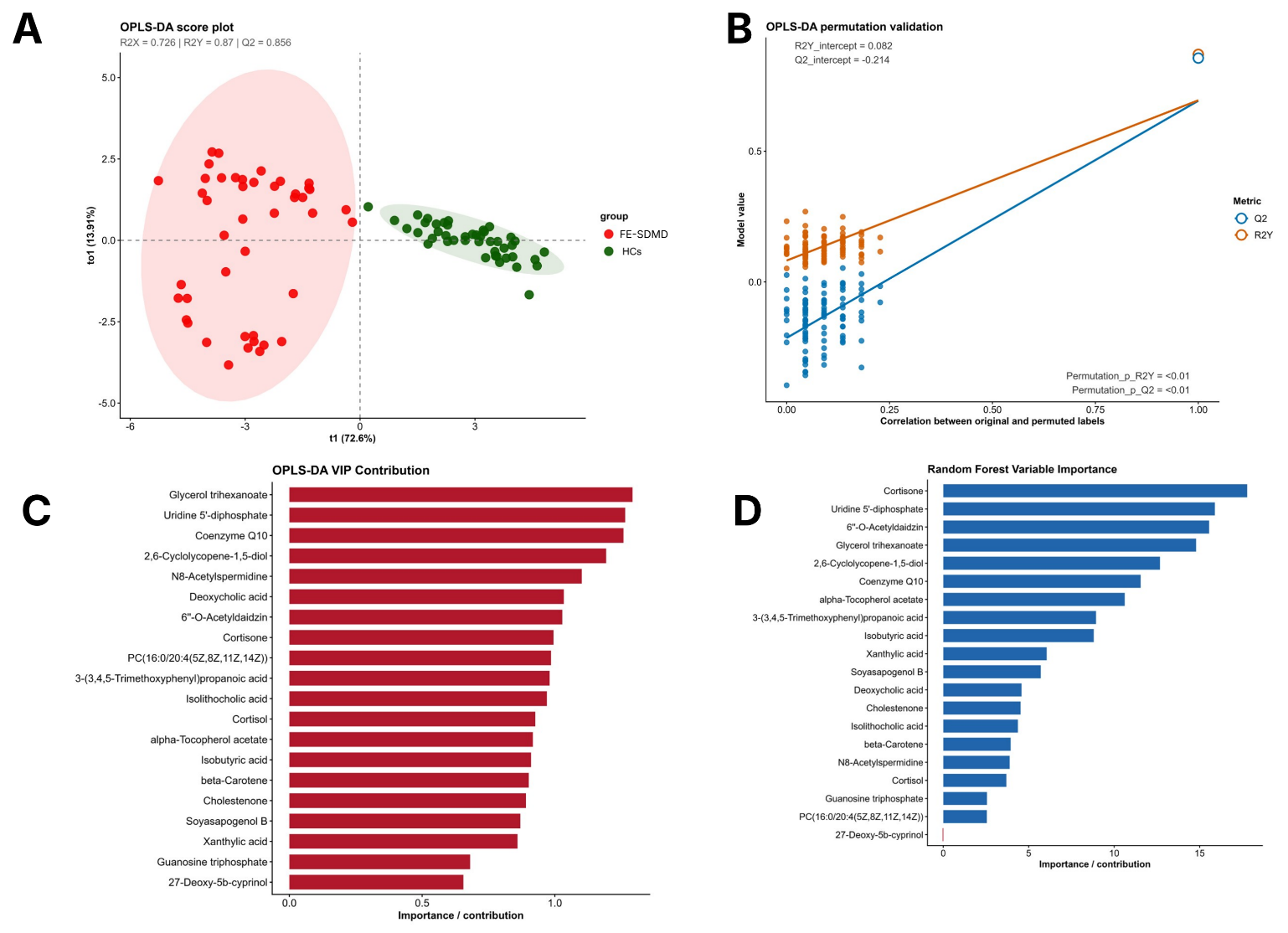

Supplementary Figure 6

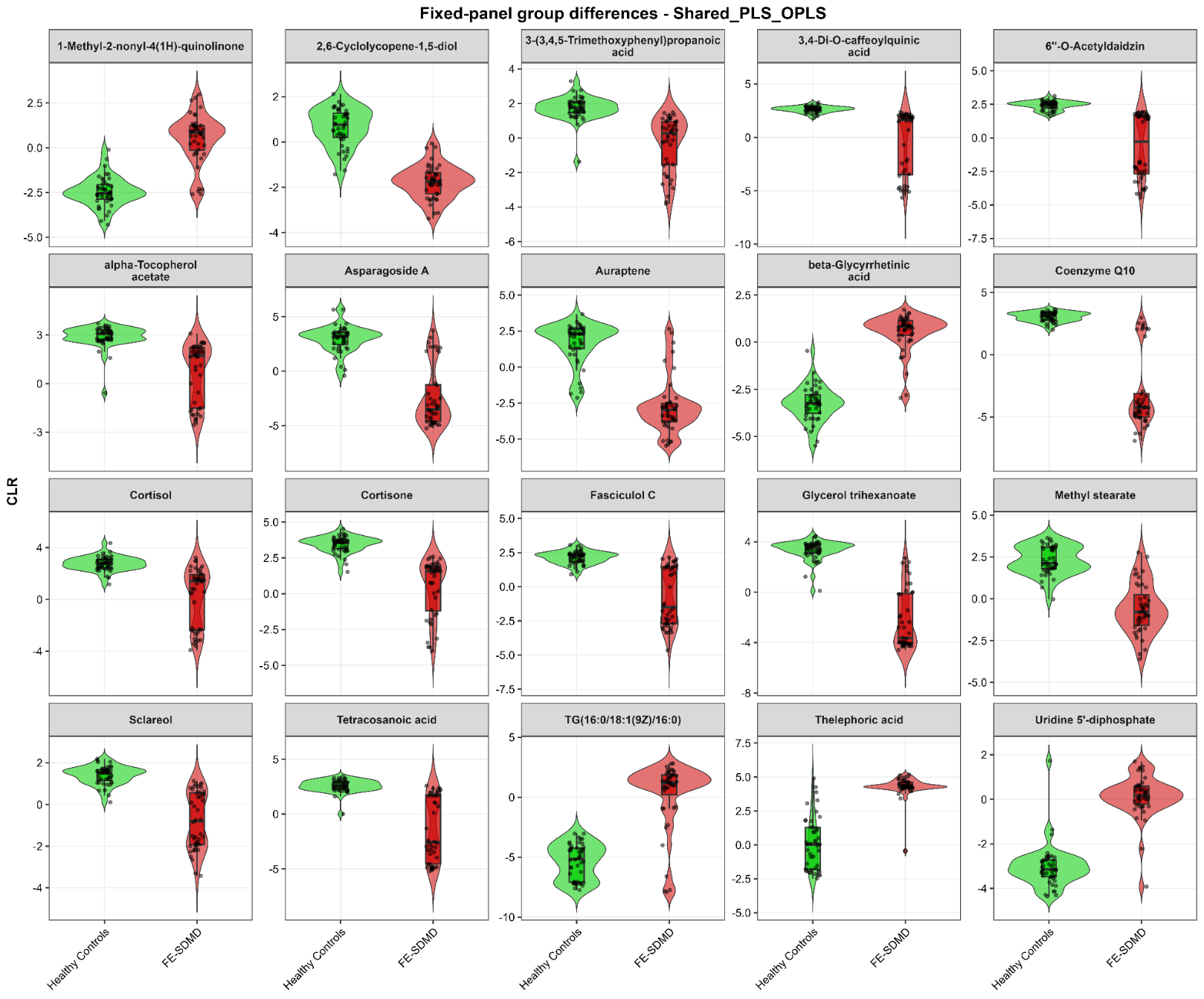

**Supplementary Figure 1.** The OPLS-DA model based on all metabolites (Supplementary Figure 1A) showed good discrimination, with adequate nested cross-validated performance. The selected metabolite panel (Supplementary Figure 1B) retained the same nested CV AUC and sensitivity.

**Supplementary Figure 2. Sparse partial least squares-discriminant analysis (sPLS-DA) of the training dataset.** **(A)** sPLS-DA score plot showing clear separation between patients with first-episode simple dysmood disorder (FE-SDMD; red) and healthy controls (HC; green). The first component explained 74.17% of the discriminative variance, and the model achieved a cross-validated area under the receiver operating characteristic curve (CV-AUC) of 0.999 and a balanced accuracy of 0.977. Shaded ellipses represent the 95% confidence regions. **(B)** Permutation validation (500 permutations) demonstrating that the observed CV-AUC (red vertical line) was significantly greater than expected by chance (p = 0.00498), indicating excellent model robustness and minimal overfitting. **(C)** Selection-frequency plot showing the proportion of repeated sPLS-DA models in which each metabolite was retained. Metabolites with the highest selection frequencies represent the most stable and reproducible discriminative biomarkers and were integrated with complementary machine learning and regression-based feature-selection methods to derive the final consensus biomarker panel.

**Supplementary Figure 3. Feature importance rankings derived from recursive feature elimination (RFE) and Boruta analyses.** **(A)** Mean feature importance scores obtained from recursive feature elimination (RFE), ranking metabolites according to their contribution to classification performance during iterative variable selection. **(B)** Mean Boruta importance scores for metabolites confirmed as informative by comparison with randomized shadow features. The marked overlap between the highest-ranked metabolites identified by both approaches demonstrates the robustness and stability of feature selection across independent machine learning algorithms. These results were integrated with PLS-DA, OPLS-DA, sparse PLS-DA, Elastic Net, bootstrap, and stability-selection analyses to construct the final consensus biomarker panel.

**Supplementary Figure 4.** Hierarchical clustering heatmap of the selected biomarker panel (n=51) (Supplementary Figure 4A). Columns represent individual participants and rows represent metabolites. Metabolite abundances are displayed as standardized (z) scores, with blue indicating lower and red indicating higher relative abundance. The upper annotation bar denotes diagnostic group (green, healthy controls; red, first-episode simple dysmood disorder [FE-SDMD]). Supplementary Figure 4B shows the PCA score plot of this selected biomarker panel. The PCA model was developed in the training dataset and the independent testing samples were subsequently projected into the training-derived principal component space without model refitting. Triangles denote training samples and circles denote testing samples.

**Supplementary Figure 5. Validation and interpretation of the final OPLS-DA model comprising 20 selected metabolites** showing clear separation between patients with first-episode simple dysmood disorder (FE-SDMD; red) and healthy controls (HC; green). The model demonstrated good performance (R²X = 0.726, R²Y = 0.870, Q² = 0.856), with shaded ellipses indicating the 95% confidence regions. Panel B: permutation validation (500 permutations) showing that both R²Y and Q² significantly exceeded values obtained under random class assignment (both permutation p < 0.01). The low R²Y intercept (0.082) and negative Q² intercept (–0.214) indicate that the model is robust and not overfitted. Panel C: shows the Variable Importance in Projection (VIP) ranking as obtained by OPLS-DA; Panel D: variable importance as obtained by random forest.

**Supplementary Figure 6. Individual distributions of the final consensus biomarker panel in healthy controls and patients with first-episode simple dysmood disorder** **(FE-SDMD).** Violin plots show the distributions of centered log-ratio (CLR)-transformed metabolite abundances in healthy controls (green) and FE-SDMD patients (red). Individual observations are overlaid as points, and embedded boxplots indicate the median and interquartile range. The displayed metabolites constitute the final consensus biomarker panel selected through the integration of PLS-DA, OPLS-DA, sparse PLS-DA, Elastic Net, recursive feature elimination, stability selection, bootstrap selection, and Random Forest/Boruta analyses. The figure demonstrates consistent and substantial differences in metabolite abundances between FE-SDMD and healthy controls, with limited overlap between groups, supporting the robustness of the selected biomarker panel.
